# Bactogram: Spatial Analysis of Bacterial Colonization in Epidermal Wounds

**DOI:** 10.1101/2024.04.18.24305961

**Authors:** Karl Wallblom, Fredrik Forsberg, Sigrid Lundgren, Jane Fisher, José Cardoso, Ganna Petruk, Ann-Charlotte Strömdahl, Karim Saleh, Manoj Puthia, Artur Schmidtchen

## Abstract

Skin barrier damage and subsequent development of harmful microbiota contribute to conditions such as wound infections, atopic dermatitis, and chronic wounds, which impact millions of people globally and pose a significant economic burden on healthcare systems. Established microbial sampling methods, such as swabs and tissue biopsies, provide limited information on the spatial distribution of bacteria. We here describe a new method that produces a visual map of the distribution of cultivable bacteria, denoted “Bactogram”, across the whole wound and surrounding skin, suitable for image-based quantification. As part of an exploratory endpoint in a clinical trial (NCT05378997) we applied the Bactogram method to 48 suction blister wounds in 24 healthy volunteers. Bacteria developed in all wounds, predominantly on the skin under the dressing and near wound edges. Two quantification methods, based on visual scoring and image analysis, demonstrated high inter-, and intra-rater agreement and were used to characterize bacterial re-colonization during epidermal wound healing. We also demonstrated proof of concept that the method can be used with chromogenic agar to enable spatial identification of pathogenic bacterial species, such as *Staphylococcus aureus*. In conclusion, this study introduces a simple method for sampling bacteria over large areas and generating a bacterial map that can identify spatial variations in bacterial composition and abundance in skin and wound conditions.

## 1. Introduction

The skin serves multiple vital functions such as impeding water loss ^1^, preventing infiltration of harmful substances ^1^, and hosting beneficial microbial communities ^2–4^ while simultaneously safeguarding against excessive levels of pathogenic microorganisms ^2, 5^. However, when the healthy skin barrier is compromised, for instance by wounding or inflammation, this delicate equilibrium is disrupted, creating an opportunity for potentially harmful bacteria to colonize ^6^. This is a known pathogenic mechanism in a panorama of debilitating conditions, including wound infections ^7^, atopic dermatitis ^8^, and chronic wounds ^9^. Understanding how the microbiota develops following skin barrier disruptions is a significant step toward prevention and treatment of these conditions.

In recent years, the ability to identify bacterial species in skin and wound samples by genomic-based techniques has advanced significantly, enabling the analysis of the entire microbial flora rather than just the dominant species ^10, 11^. However, current sampling techniques, including swabbing and tissue biopsies, restrict our ability to investigate spatial differences as they typically sample a specific area and offer no information regarding spatial variations of microbiota within the sampled region. The overwhelming majority of the available literature focuses on bacterial samples taken from the wound center ^7, 12^. However, emerging evidence indicates that bacteria are also present in substantial quantities at the wound edges in acute ^13^ and chronic wounds ^14^. The wound edges are vital for wound healing ^15, 16^, calling for further studies investigating their bacterial levels, as generally high levels may warrant additional bacterial management in clinical routines.

This study introduces a new method to address existing limitations in studying the spatial distribution of bacteria *in vivo*. At its core, this method involves applying moist filter paper to the subject’s wound and skin to collect bacteria, which is then directly transferred to culturing plates. The entire procedure produces a visual map of the distribution of cultivable bacteria, denoted “Bactogram”. A Bactogram covers an area of approximately 120 cm^2^ and is highly suitable for image-based quantification.

In this study, we aimed to evaluate the applicability of the Bactogram method and examine the spatial development of bacteria in the context of acute wounding, while also comparing the results with more established microbial sampling techniques like swabbing. We evaluated the method in standard wound healing conditions by applying it to 48 placebo-treated suction blister wounds in healthy volunteers in a clinical trial investigating the safety of an antimicrobial and immunomodulating wound gel ^17^. Suction blister wounds are highly reproducible in size and depth and heal without scarring ^18^, and therefore well-suited for investigating the dynamics of microbial re-colonization following a defined skin barrier disruption.

Following the same wounds over 11 days allowed us to describe and quantify the spatial colonization of cultivable bacteria during wound healing. We revealed that, while the overall spatial pattern of re-colonization after wounding was relatively consistent, substantial variation existed between different wounds. The skin beneath the dressing rather than the actual wound, was identified to be most commonly and densely colonized.

## 2. Material and methods

### 2.1 Samples

The samples, data, and images used in this study were collected and processed as part of an exploratory endpoint in a randomized-controlled clinical trial evaluating the safety of topical application of a TCP-25 wound gel on suction blisters induced on 24 healthy volunteers (clincaltrials.gov: NCT05378997). The study was conducted in accordance with the Declaration of Helsinki and was approved by Etikprövningsmyndigheten (Swedish Ethics committee) and Läkemedelsverket (Swedish Medical Products Agency). Written informed consent was collected from all included subjects. A detailed explanation of the clinical trial design and the other procedures within the trial not directly relevant to the Bactogram method can be found in the published clinical trial protocol ^17^.

### 2.2 Method Details

#### 2.2.1 Wound induction and wound care

Suction blister wounds were induced on 24 healthy volunteers in the clinical trial described above ^17^. Prior to wounding, hair at the wound sites was shaved, and each site was wiped with an ethanol-soaked gauze. The blisters were made using the Model NP-4 (Electronic Diversities, USA) suctioning device ^18^. Two 10 mm diameter blister wounds were created on the medial aspect of each thigh, with a 6 cm distance between them. After the blisters formed, the blister roof was excised using sterile forceps and scissors.

In this study, we used only the control (placebo-treated) wounds from each subject (48 control wounds) to verify and describe the Bactogram method. The placebo treatment was a topical application of 0.15 mL of a proprietary hydrogel (Pharma-Skan ApS, Denmark) with 1.4 % hydroxyethyl-cellulose (Ashland, USA) at pH 7.0 ^17^. The wounds were dressed using a 2 × 2 cm polyurethane foam primary dressing (Mepilex transfer, Mölnlycke Health Care, Sweden) and further covered with a secondary dressing (Tegaderm, 3M, USA), a secondary protective layer consisting of overlapping gauze swabs, and another tertiary Tegaderm dressing. Dressings were applied after wounding on day 1 and changed during study visits on days 2, 3, 5, and 8. On day 11, only gauze weave was applied as primary protection, covered by a Tegaderm film.

Further details on other study procedures not directly relevant to the present study can be found in the published protocol ^17^.

#### 2.2.2 Bactogram collection

To create bacterial replicas of the subject’s wound and cultivable skin bacteria, sterile Munktell filter paper (size 125mm, Grade 3, Ahlstrom-Munksjo, Sweden) was used. To ensure sterility, the filter papers were autoclaved in autoclave bags (SPS Medical, France) using the Instrument/Textiles 121°C standard program on an HS33 autoclave (Getinge, Sweden). To keep track of the orientation of the Bactogram, one notch was cut at 12 o’clock and two notches at 3 o’clock for each filter paper (Figure 1A).

**Figure 1:**
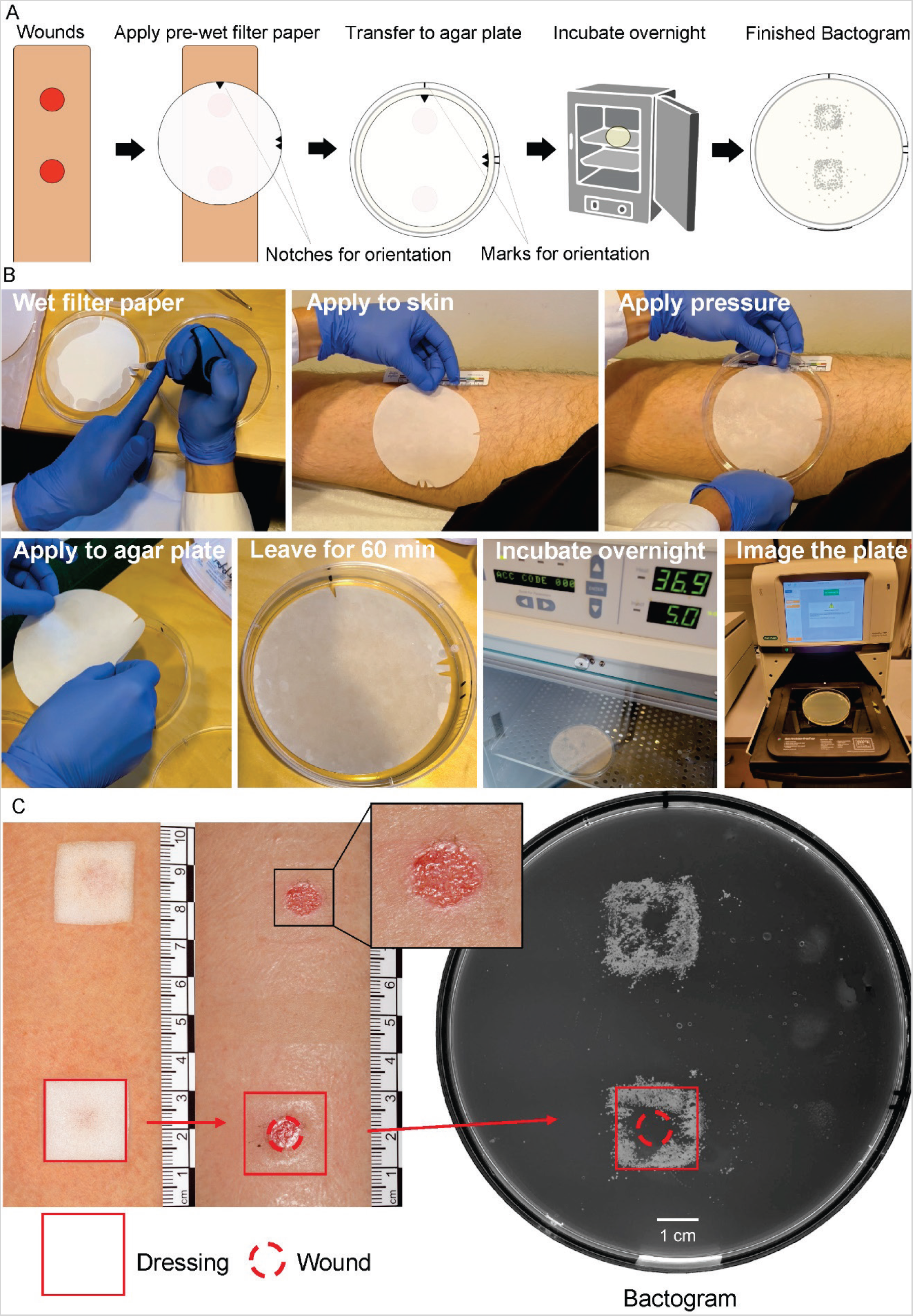
Overview of the method for Bactogram acquisition. (A) Illustration demonstrating the workflow for acquiring Bactograms. (B) Images of the most important steps in the Bactogram workflow. Step 1, pre-wet filter paper in a clean Petri dish using 2 ml of sterile PBS. Step 2, apply the filter paper to the skin. In this study, one notch was aligned with the proximal wound (wound closest to the groin) and the side with two notches to the subject’s left, to get a consistent orientation. Apply pressure using the clean lid of the Petri dish. Step 3, remove the filter paper from the skin and transfer the filter paper skin-side down to the agar plate. Ensure the notches or markings align with their corresponding marks on the agar plate. Step 4, let the filter paper sit on the agar plate at room temperature for 60 minutes before discarding the filter paper. Step 5, incubate the agar plate overnight at 37° C, 5% CO_2_. Step 6, image the Bactogram using an appropriate imaging system such as the Chemidoc MP imaging system (Bio-Rad, USA). (C) Images of representative suction blister wounds and dressings and the spatial relation between the dressing, wound, and resulting Bactogram.

Approximately 10 minutes before the Bactograms were collected, the filter paper was placed in an empty 15 cm sterile plastic petri dish, and 2 ml of sterile phosphate buffered saline (PBS, pH 7.4), was added using a pipette to pre-soak the filter paper. Problems with “smearing” of the bacterial colonies on the agar plates were noted for the initial Bactograms due to excessive moisture on the agar plates (Supplementary Figure 1). The problem was solved by drying the plate by applying a dry filter paper (size 125mm, Grade 5, Ahlstrom-Munksjo, Sweden) on the agar plate which is discarded after absorbing the moisture (approximately 5 min).

After removal of the dressing from the wounds at each study visit, but before swabbing and the application of the placebo gel, the pre-soaked filter paper was gently placed on the subject’s leg, using forceps. The filter paper was placed so that it covered both wounds and so that the side with one notch was aligned with the proximal wound (wound closest to the groin) and the side with two notches to the subject’s left (Figure 1B). The lid of the Petri dish was used to press the filter paper lightly against the leg for 1 minute using a slight rocking motion to ensure equal contact of wounds with the filter paper.

Afterwards, the filter paper was removed from the skin using forceps and placed onto a 15 cm Todd Hewitt agar plate, skin-side down. To preserve the orientation of the wounds in the Bactogram, the side on the filter paper with two notches was aligned with two pen marks on the plate, and the side with one notch was aligned with one pen mark on the plate (Figure 1B). The plates with the filter paper were then incubated for 1-hour at room temperature.

After 1 hour of incubation at room temperature, the filter paper was discarded and the agar plates were incubated at 37 °C in 5% CO_2_ overnight. An example of a Bactogram is depicted in Figure 1C.

#### 2.2.3 Image acquisition and Bactogram quantification

After the overnight incubation, the agar plates were photographed using a ChemiDoc MP Imaging System and Image Lab imaging software (Bio-Rad, USA). The Stain Free Blot imaging application was used with a manual exposure time of 0.386 s and a size of 20 cm × 16 cm, yielding grey-scale images of all Bactograms. The colors were then inverted using the Chemidoc MP Imaging System to make bacterial colonies brighter than the agar, to enhance visualization.

We developed two different methods to quantify bacterial coverage in the Bactogram images. One utilizing a manual visual scoring system and one utilizing image analysis via ImageJ Fiji versions 2.5.0 - 2.11.0 ^20^. Two different areas of the Bactogram, corresponding to areas of clinical and scientific interest, were quantified independently, namely the “Wound” and “Dressing” areas. The wound area was defined as the area where the original blister was made. This area stayed the same, even if the wound healed or closed. The dressing area was defined as the area under the dressing, other than the wound area.

#### 2.2.4 Visual scoring system

A visual Bactogram scoring system was developed using a 5-point scale, with 0 being the lowest score (corresponding to no visible bacterial coverage) and 4 being the highest score (corresponding to near complete coverage by bacteria). This scale was chosen as it was deemed to have enough granularity to differentiate between different levels of bacterial coverage while still being easily differentiated by raters.

The scoring was done by three independent raters, all working at the Department. The identity of the wounds (subject nr, leg, and day) was not indicated in the images. However, the images were only partially masked as the images were provided in order by the subject number, but the raters were not aware of this. A detailed description of the scoring system and instructions were given to the raters in advance (Supplementary Data 1). This document includes an explanation of how to score bacterial coverage in three areas (wound, dressing, and skin), as well as visual aids for each grade. However, the skin area was not quantified in this study. Nonetheless, we include a description of how such a quantification could be done using the visual scoring system.

The raters’ agreement was calculated using Krippendorff’s alpha to determine the inter-rater reliability. One rater also rescored 50 randomly selected images, excluding day 1, to investigate the scoring system’s intra-rater reliability, using Krippendorff’s alpha.

#### 2.2.5 Computer-assisted quantification

An alternative method of quantification was created using image analysis through ImageJ (Fiji versions 2.5.0 - 2.11.0) to calculate the area covered by bacterial colonies more objectively and obtain continuous rather than ordinal data.

Using ImageJ, all Bactogram images were converted into 8-bit images using the built-in 8-bit conversion function to enable the use of the threshold function. A threshold value for each Bactogram image was chosen manually using the Threshold dialogue box. The upper threshold values were kept at 255 while the lower values were manually adjusted until the bright edges of the petri dish in each image were of comparable apparent intensity, with minimal image artifacts. Once the threshold was selected, ImageJ generated a binary black and white image to enable quantification of the area covered by bacteria, now displayed in white.

To quantify the bacterial coverage in the dressing and wound area, we first calculated the exact size of the area on the Bactogram images corresponding to the 2 × 2 cm area under the primary Mepilex dressing. To do so, we first imaged a Mepilex dressing laid out on an agar plate using the same settings as for the agar plates described above. The image used to determine the size of a Mepilex cut-out can be found in Supplementary Data 2. The area of the Mepilex dressing was measured to be 250 × 250 pixels in ImageJ. A macro within the ImageJ program (Supplementary Data 3) was then used to quickly recreate this square area and prompt the user to position the square area in the region of interest on the Bactogram image (Supplementary Figure 2). The position was determined by following the decision tree in Supplementary Figure 3. Once the square area was positioned correctly, the user would click the prompt. This activated the measure command in ImageJ, which was set to measure the area fraction and integrated density, resulting in a measurement of the total intensity and % bacteria coverage within the selected area, giving us the bacterial coverage of the dressing and wound area combined. This process was then repeated for each Bactogram image.

A similar method was employed to quantify bacterial coverage in the wound area. The blister wounds had a diameter of 1 cm, corresponding to a diameter of 125 pixels, as the 2 × 2 cm dressing previously was shown to measure 250 × 250 pixels in the Bactogram image. A macro within the ImageJ program was used to quickly recreate the circle and prompt the user to position the circle at the region of interest on the Bactogram image corresponding to the wound area, similarly as described earlier for the dressing area. Once the circle was positioned correctly, the user would click the prompt to activate the measure command. For calculating bacterial coverage in the area swabbed, used in the comparison with conventional quantitative bacterial count, we modified this method. Specifically, we expanded the circle’s diameter by 40% to account for the swabbing protocol in the clinical trial, which involved swabbing the wound along with an additional 2 mm margin on each side.

The dressing area was earlier defined as the area under the dressing, other than the wound area for the visual scoring system. To replicate this measurement for the computer-assisted method, we subtracted the results of the wound area from that of the combined dressing and wound area and then divided it by the whole dressing size minus the wound size (Eq 1).

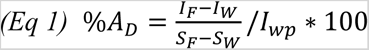

%A_D_: % Area covered dressing, I_F_: Intensity full area (wound and dressing), I_W_: Intensity wound area, S_F_: Size full area (62500 pixels), S_W_: Size wound area (12281 pixels), I_wp_: Intensity of a white pixel (255).

To evaluate the reliability of the method, all computer-assisted quantifications were performed two additional times. One additional time by the same person who performed the original quantification to check the intra-rater reliability, and one time by a second person to check the inter-rater reliability. Only the first measurement was used in further statistical analyses.

#### 2.2.6 Heat map of bacterial presence

On each binary masked Bactogram image described in the above section, we cropped the images to include only an area of 350 × 350 pixels, centered around the area under the Mepilex dressing. The cropped images were grouped by study day. Images from the same day were then layered on top of each other, and the average intensity of each pixel in the layered image was calculated, resulting in a bacterial coverage heatmap, which was then visualized by applying a colormap. This was performed using a python script, found in Supplementary Data 4.

#### 2.2.7 Colony forming unit count

As part of the clinical trial, CFU counting of the swab and dressing samples was done previously and reported by Lundgren et al.^19^ In short, swabs were diluted in 500 µl of PBS, while dressings were diluted in 2 ml of sterile Tris buffer. Both the swab and dressing fluid samples were subsequently diluted in sterile PBS to produce seven sequential 10-fold dilutions, ranging from 10× to 10^7×. From each dilution, including the undiluted sample, six individual 10-μL drops were placed on agar plates. These plates were then incubated at 37°C with 5% CO2 overnight. The following day, colonies on the plates were counted and documented. To express the results in bacterial density (CFU/cm^2^), we multiplied the CFU/ml results by the sample volume and then divided by the collection area specific to each method, 1.5 cm^2^ for the swab and 4 cm^2^ for the dressing.

#### 2.2.8 Chromogenic agar

Using the Bactogram method together with chromogenic agar would extend the method’s possibilities by enabling spatial identification of different bacterial species. As a pilot study, a double Bactogram was made by making a second bacterial imprint on a chromogenic agar using the same filter paper and method as described above, after having done the usual bacterial imprint on the non-chromogenic Todd Hewitt agar. This was performed on study day 3 and 8 on the last 16 participants. The time points day 3 and 8 were chosen as MALDI-TOF mass spectrometry analysis was performed on these days as described by Lundgren et al.^19^ We chose a chromogenic agar (CHROMagar Staph aureus, France) developed to specifically identify *S. aureus* as purple/pink colonies ^21^. This choice was based on the prevalent role of *S. aureus* as the principal bacteria causing acute wound infections ^22^.

### 2.3 Statistical analysis

All statistical analyses and graphs were done in GraphPad Prism (version 9) or R Studio. All the details of the analyses can be found in the figure legends and results, including the statistical test used and sample size for analysis.

The visual scoring system generated ordinal data with whole values between 0 – 4, so they are presented as the median and interquartile range. Computer-assisted wound total intensity values generated continuous data with possible values as factors of 255. All calculated computer-assisted % area coverages generated continuous data, rounded to 3 decimal digits, within the range 0 to 100.

Krippendorff’s alpha was calculated for correlations with ordinal data using the “ordinal” method of the “kripp.alpha” function in the “irr” package in R as described by Jacob Long ^23^. Confidence intervals for Krippendorff’s alpha were calculated with a 10000 repeat bootstrapping using the “boot” function in the “boot” package in R.

For correlations of continuous data, the two-way, mixed, absolute, intra-class correlation coefficients and their respective confidence intervals were calculated and chosen according to the method described by Ko et al. ^24^ and Haghayegh et al.^25^.

Both quantification methods were compared against each other and against the corresponding CFU/cm^2^ values. This was done by correlating the results from each quantification method using a nonparametric Spearman correlation. The nonparametric Spearman correlation method was chosen as the visual scoring results are ordinal and as none of the computer-assisted quantification data were found to be normally distributed.

### 2.4 Resource availability

The generated pictures of the Bactograms will be available on reasonable request, after the publication of the treated wound data in a separate publication, since the original images inevitably include both treated and untreated wounds. The code used for the ImageJ macros can be found in Supplementary Data 3. The Python code for creating spatial heat maps can be found in Supplementary Data 4. Further information or instructions required to reanalyze the data reported in this paper are available from the lead contact upon reasonable request.

### 2.5 Additional resources

The clinical trial is registered at ClinicalTrial.gov (NCT05378997), URL: https://clinicaltrials.gov/ct2/show/NCT05378997. The full details of the clinical trial protocol are published in Lundgren et al. ^17^.

## 3. Results

### 3.1 Bactogram collection

To obtain the Bactograms, sterile and pre-wetted filter paper was pressed against wounds and their surrounding skin to collect bacteria with retained spatial distribution and further processed (Figure 1A and B), see method part for further details. To investigate the spatial development of bacteria in and around the wound over time, this method was applied to all 24 study participants at six time points post-wounding, generating two Bactograms for each participant, one per leg.

During the first three days of the first group of eight subjects, problems with “smearing” of the bacterial colonies on the agar plates were noted (Supplementary Figure 1). The cause of this was identified to be excessive wetness, due to condensation, on the agar plates. To solve this problem, a dry filter paper was first applied to each agar plate to dry excess moisture and then discarded. The images with smearing were excluded from all analyses, but a sensitivity analysis that includes images with smearing is provided in Supplementary Figure 4.

### 3.2 Visual evaluation of Bactograms

To enable visualization of the Bactograms over time, grayscale images were acquired using a Chemidoc MP Imaging System (Bio-Rad, USA). In Figure 1C, one such image is shown together with a clarification of the spatial correspondence between the wound, its dressing, and the corresponding Bactogram. By looking at the Bactogram images in chronological order, the spatial development of bacteria around the suction blister wounds is evident. The development in five different representative wounds is shown in Figure 2A. The Bactograms exhibited a variable increase in bacteria under the dressing on days 3-5, which typically persisted until day 11 when all wounds were completely healed ^26^.

**Figure 2:**
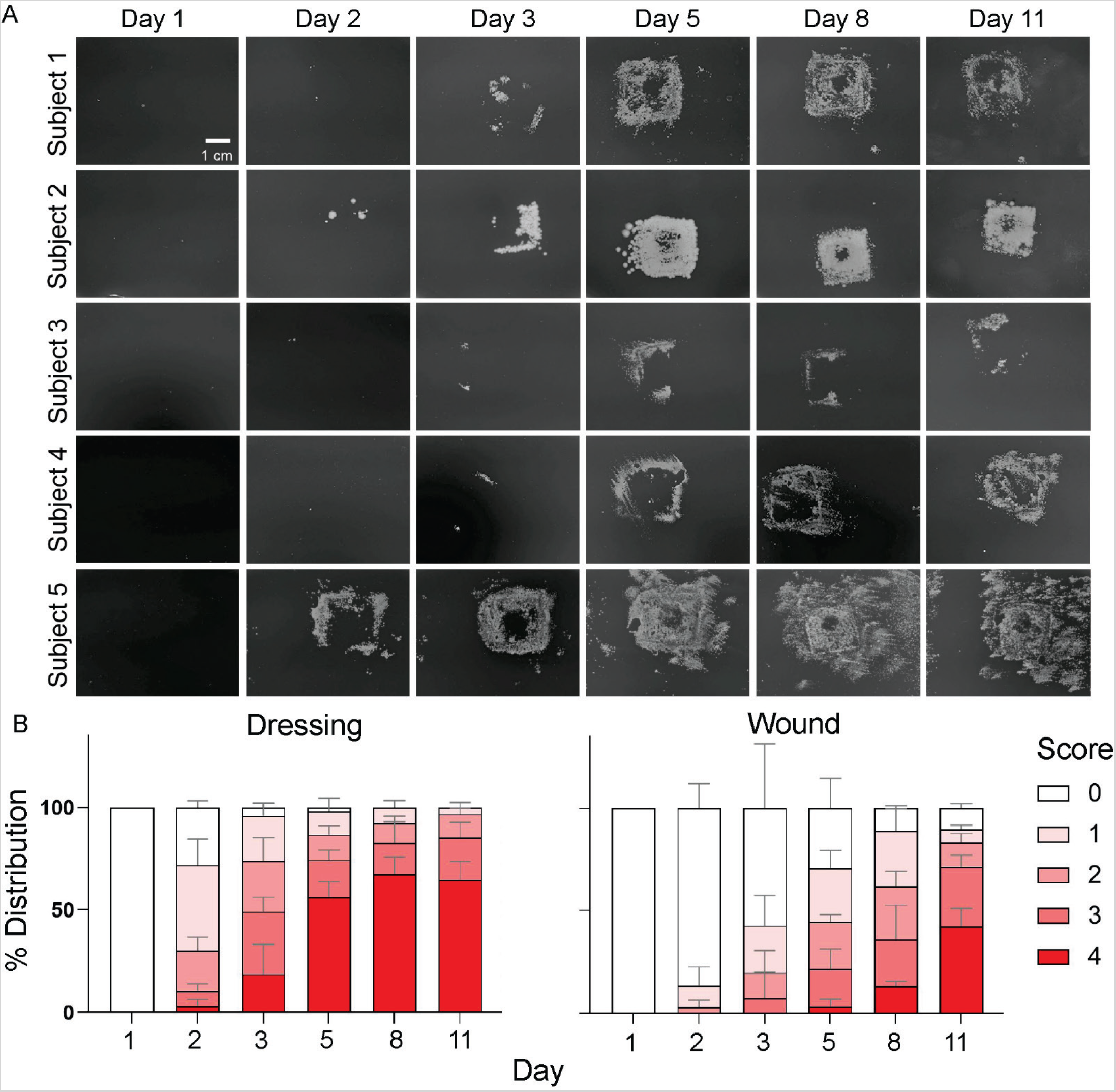
Bacterial development during wound healing visualized by the Bactogram method. (A) Visual appearance of Bactograms collected at each study visit from 5 subjects, representative of the typical Bactogram appearance and development during the study. (B) Proportional stacked bar graph showing the proportion of wounds given each score using the visual scoring system for the dressing and wound areas. The two areas corresponding to the dressing and wound, explained in Figure 1C, were scored on a 5-grade scale between 0 – 4, where a score of 0 indicates no bacterial colonies in that area, and a score of 4 indicates that the area is completely covered by bacteria. Each wound was scored by three independent raters given the same instructions. The sample size is 24 wounds for each time point except for days 1, 2, and 3, where the sample size is 16 due to the exclusion of smeared Bactograms from the first 8 subjects at these time points. The bars show the mean proportion of each score and the error bars show the standard deviation of the proportion of each score by the three different raters.

To quantify the Bactograms we established a visual scoring system. This system is based on a visual evaluation of the amount of bacterial colonies in the wound and dressing area (Figure 1C) on a 5-grade ranging from 0 (no bacterial colonies) to 4 (near complete coverage with bacterial colonies) (Supplementary Data 1). Three independent raters used this scoring system to quantify the amount of bacterial colonization in the wound and dressing area of all wounds. The complete score data from all three raters is provided in Supplementary Data 5.

To investigate the spatial development of the bacteria after wounding, we calculated the proportion of each score for each day in the dressing and wound areas (Figure 2B). Directly after wounding (Day 1), the dressing and wound areas showed no bacterial coverage. Overall, bacterial coverage in the wound area generally increased progressively until the end of the study (day 11). In contrast, bacterial coverage in the dressing area generally peaked on Day 5/8 and remained high until Day 11 (Figure 2B).

To investigate this visual scoring method’s reliability, we calculated inter- and intra-rater reliability using Krippendorff’s alpha (Table 1), revealing high intra-rater reliability for both the wound (0.880) and dressing areas (0.936). Inter-rater reliability was moderately lower yet acceptable for both regions (Wound: 0.792; Dressing: 0.828). 95% confidence intervals and probabilities of failing to reach 0.667 were calculated using bootstrapping with 10000 repeats. All the rater reliabilities exceeded 0.667 in all bootstrapping repeats.

**Table 1.** Reliability of the visual Bactogram scoring system as indicated by Krippendorff’s alpha.

|  | Krippendorff's alpha, 3<br>raters inter-rater reliability<br>[95% confidence interval]<br>{Probability of < 0.667} | Krippendorff's alpha, one<br>rater, intra-rater reliability<br>[95% confidence interval]<br>{Probability of < 0.667} |
| --- | --- | --- |
| Wound | 0.792 [0.733 - 0.837] {0} | 0.880 [0.772 - 0.940] {0} |
| Dressing | 0.828 [0.776 - 0.869] {0} | 0.936 [0.848 - 0.991] {0} |

The visual scoring system, while facilitating the quantification of Bactograms, presents several challenges. A notable concern is the time-consuming nature of the manual scoring process, with the 288 wounds taking approximately four hours for a rater to assess. Moreover, the inherent subjectivity in visual evaluations introduces potential biases and the risk of human errors. Additionally, while we observed relatively high inter- and intra-rater reliability in this study, it’s worth noting that our raters had prior experience with Bactograms resulting in a higher reliability than can be expected for less familiar raters.

### 3.3 Computer-assisted quantitative analysis of Bactograms

In an attempt to overcome certain limitations of the visual scoring system such as its time-consuming nature, subjectivity, and modest inter-rater reliability, we developed a computer-assisted image analysis method using ImageJ ^20^. This method offered a way to quantify the extent of bacterial coverage in the area of interest expressed as a continuous variable ranging from 0-100, as opposed to the four-grade categorical visual scoring system. The workflow for this method is outlined in Figure 3A. Briefly, this method applies a binary mask with individual threshold values to each grayscale Bactogram image, rendering binary Bactograms that portray areas of bacterial growth in white and non-growth areas in black (Figure 3A). The bacterial coverage is then measured by manually selecting the wound and dressing area in the binary Bactogram images using an ImageJ script. All measurements from the binary images are available in Supplementary Data 5.

**Figure 3:**
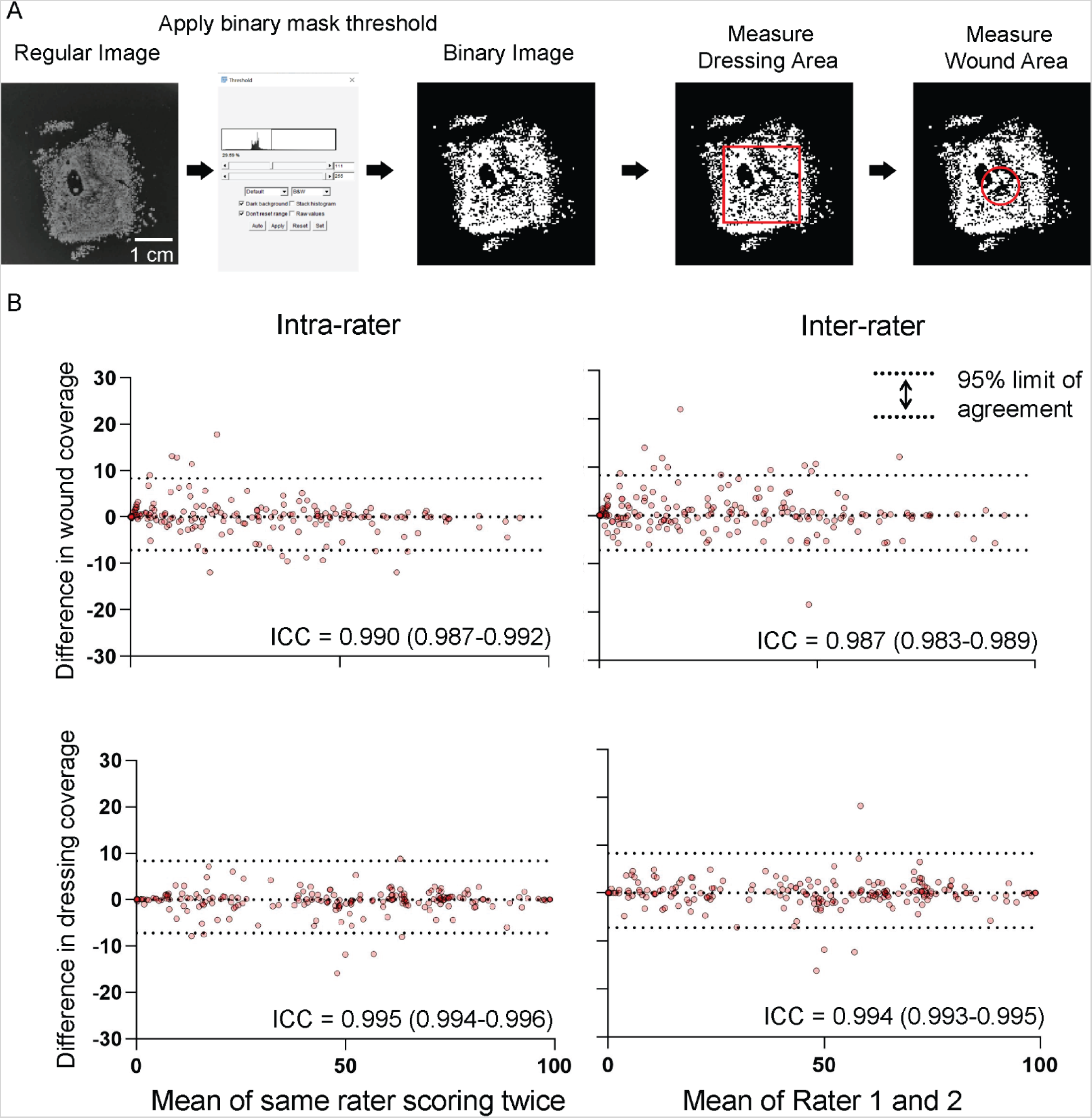
Computer-assisted image analysis of Bactograms. (A) Illustration of the general workflow for binarization and quantification of Bactogram images. The Bactogram images were converted to a binary representation indicating bacterial presence (white) or no bacterial presence (black) at each pixel using the threshold function in ImageJ. Bacterial coverage in the dressing and wound areas was then quantified by calculating the percentage of white pixels in the dressing and wound area. (B) Bland-Altman plots visualizing differences in the bacterial coverage measurements by the same rater twice (intra-rater) and two separate raters (inter-rater). The day 1 time point is not included in the graph as almost all Bactograms had zero bacterial coverage at that time point, which would unduly skew the graph and correlation. The total sample size is therefore 104. Each data point is visualized by a semi-transparent red dot, such that a redder color indicates overlapping dots. The width of the 95% agreement indicates the robustness of the scoring method. The method’s robustness is further described by the Intra-Class Correlations (ICC), calculated using two-way, mixed, absolute, intra-class correlation, yielding a score for each correlation, with all ICC > 0.980. The 95% confidence interval of the ICC is shown in parentheses.

To investigate the reliability of the method, intra- and inter-rater reliability were determined. The differences between one rater scoring the same images twice as well as two raters scoring all images can be seen in Bland-Altman plots (Figure 3B). All two-way mixed absolute intraclass correlation coefficients, including the lower bound of the 95% confidence intervals, were found to be >0.98 (Figure 3B), which indicates excellent intra- and inter-rater reliability ^24^. Only the first measurement was used in further statistical analyses. The previously described visual scoring method and the computer-assisted quantification method were compared and correlated, showing a good correlation but with substantial overlap in computer-assisted values between the scoring levels (Supplementary Figure 5).

### 3.4 Investigation of bacterial growth patterns via computer-assisted image analysis

Bacterial coverage showed a similar pattern over time when quantified using the computer-assisted analysis (Figure 4A) as it did when quantified using the visual scoring system (Figure 2B). As seen previously, bacterial coverage in the wound area generally increased progressively throughout the study, while bacterial coverage in the dressing area peaked on Day 5/8, and remained at that level until Day 11 (Figure 4A). The computer-assisted method also illustrates the variations between different wounds as seen by the large interquartile range and min/max values.

**Figure 4:**
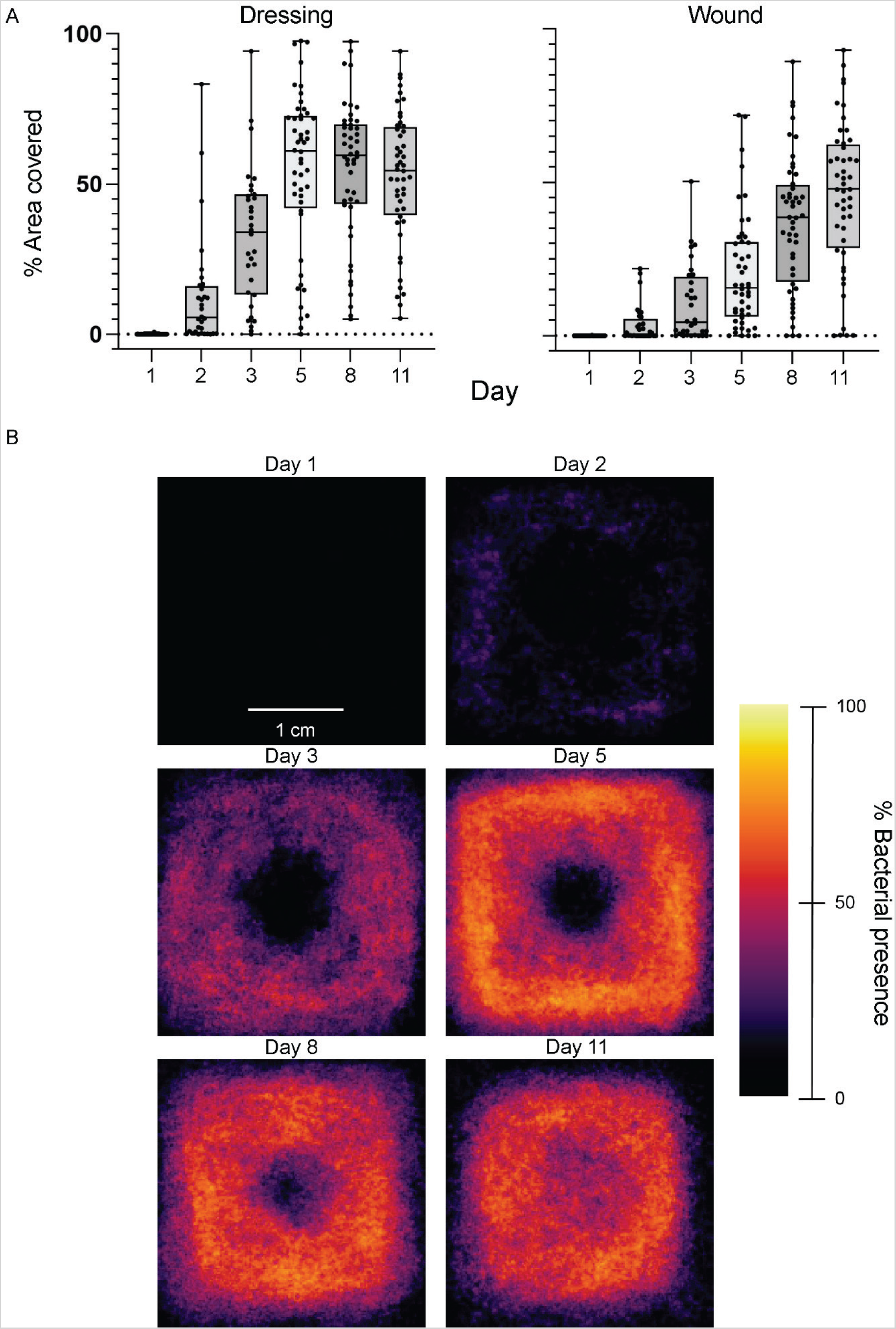
Bacterial development during wound healing quantified using computer-assisted image analysis. (A) Using our method for computer-assisted image analysis, the bacterial coverage was calculated for the wound and dressing areas of each wound. Each value is displayed using a boxplot for each time point to investigate the bacterial development over time. The boxplot shows all individual data points and indicates the median and the 25th and 75th percentile, with whiskers indicating the range (min to max). The sample size is 24 wounds for each time point except for days 1, 2, and 3 where the sample size is 16 due to the exclusion of the smeared Bactograms from the initial 8 subjects at these time points. (B) The general bacterial distribution was visualized using spatial heat maps. All Bactograms were binarized, and all images from each timepoint were superimposed. The heatmap visualizes the percentage of Bactograms having bacterial coverage at each spatial position in and around the wound. A lighter, more yellow color signifies a higher percentage of all Bactograms having bacterial coverage at that particular location. The sample size is 24 wounds for each time point except for days 1, 2, and 3 where the sample size is 16 due to the exclusion of the smeared Bactograms from the initial 8 subjects at these time points.

In addition, the binarization of the Bactogram images enabled more advanced analyses such as the creation of spatial heat maps delineating the most common locations for bacterial growth in and around the wound, for each day (Figure 4B). To create the spatial heat map, the binary picture from all wounds at each time point was overlapped and the value (0, no colony; 1, colony) at each overlapping pixel was summed (Supplementary Figure 6). Dividing the resulting sum for each of the overlapping pixels by the total number of overlapping pictures provided us with the percentage of images that had bacteria present at that specific location. This percentage was then used to color the heat map according to the color scale (Figure 4B). The resulting heat maps showed that the bacterial coverage in the wound area was generally low during the first three days. However, during the following days (Day 5 and onward) progressive bacterial colonization occurred from the edges towards the center of the wound. The skin area underneath the dressing had a higher percentage of bacterial coverage than the wound area at all time points, peaking at around day 5/8.

### 3.5 Correspondence of Bactogram quantification with conventional colony forming unit quantification

The bacterial coverage measure from the computer-assisted analysis of Bactograms was compared with bacterial density (CFU/cm^2^) measured by dividing traditional colony forming unit (CFU) counts by the area sampled. As part of the clinical trial, CFU counts were performed on swab samples collected from the wound area extended by approximately 2 mm on each side of the wound and on wound fluid extracted from the 2 × 2 cm dressings.

We hypothesized that the bacterial coverage in the dressing area and the bacterial density of the dressing fluid from the same wound should correlate with each other. We also hypothesized that the bacterial coverage in the wound area and the bacterial density of swabs from the same wound should correlate with each other. However, the area swabbed included 2 mm of the skin around the wound edge on each side of the wound and did therefore not exactly correspond to the wound area defined in the computer-assisted method. Thus, we additionally quantified bacterial coverage in an area corresponding to the actual area swabbed (the wound area extended by 2 mm on each side). Further clarification of the areas measured and how they correspond to the actual wound and Bactogram image can be seen in Figure 5A. To visualize the correlation between CFU/cm^2^ and bacterial coverage, we plotted the base-10 logarithm of the colony forming units (log CFU/cm^2^) against the percent bacterial coverage for corresponding areas (Figure 5B).

**Figure 5:**
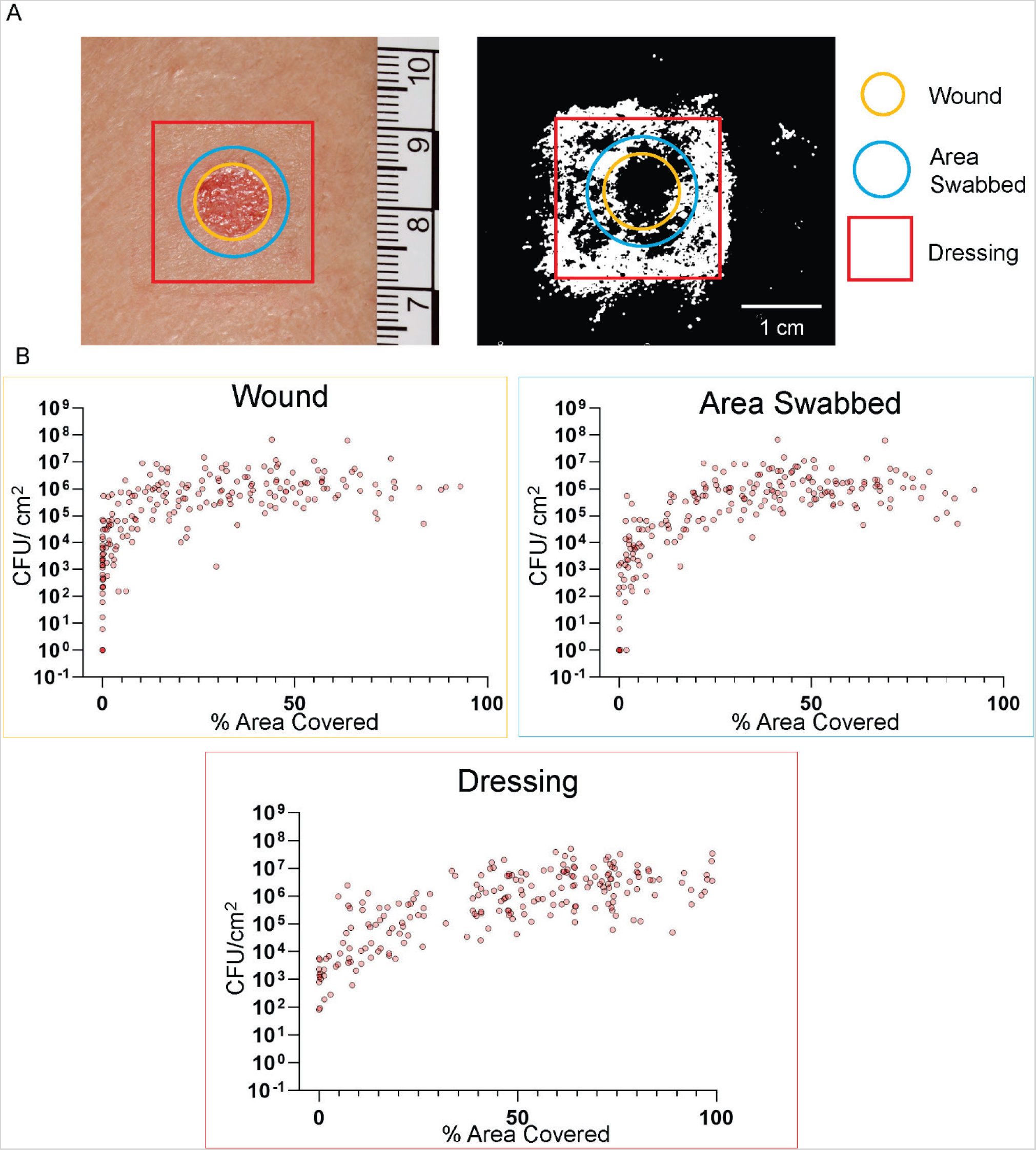
Correspondence between bacterial sampling using the Bactogram method and quantitative bacterial counts in swab and dressing fluid samples. (A) Visual representation of the spatial relation between the areas from which swab (blue) and dressing fluid (red) samples were collected and the area quantified using the computer-assisted image analysis (yellow). (B) XY-plot visualizing the correspondence of colony-forming units (CFU/cm^2^) from the swab and dressing samples with the corresponding bacterial coverage measurement calculated using computer-assisted image analysis. Data from the day 1 time point are not included in the graph as almost all values at that time point had zero CFU and bacterial coverage, which would unduly skew the graph and correlation. The total sample size is 104. Each data point is visualized by a semi-transparent red dot, such that a redder color indicates overlapping dots. The bacterial coverage in the wound area was zero or close to zero in many images. In contrast, the quantitative bacterial counts in the corresponding swab indicated a higher bacterial density, resulting in a relatively poor correlation. This effect was mitigated when the quantitative bacterial counts in the swab samples were compared with the bacterial coverage in the actual swabbed area (the wound area extended by 2 mm, blue circle).

In many Bactograms, bacterial coverage in the wound area was zero or close to zero, while the corresponding swab showed a higher bacterial density, resulting in a relatively poor correlation. This effect was mitigated when the quantitative bacterial counts in the swab samples were compared with the bacterial coverage in the actual swabbed area (the wound area extended by 2 mm). However, it should be noted that even though this modification improved the correlation, the swabbing procedure is prone to small human errors, extending or contracting the swabbed area, which will affect the results.

To calculate the correlation coefficient, we excluded Day 1 results due to a large number of samples exhibiting zero bacteria (CFU) in the swab and zero bacterial coverage on the Bactogram, which would have skewed the correlation. Spearman’s correlation comparing the computer-assisted method with the corresponding bacterial density for that wound calculated in CFU/cm^2^, resulted in r-values of 0.7124 (swab CFU/cm^2^ and wound area), 0.7105, (swab CFU/cm^2^ and swabbed area) and 0.6900 (dressing CFU/cm^2^ and dressing area). All p-values from the correlation analyses were less than 0.0001.

### 3.6 Chromogenic agar

As a proof of concept for another possible application of the Bactogram method, for some of the participants, a second bacterial imprint was made on a chromogenic agar plate with the same filter paper, after having made the usual imprint on the Todd Hewitt agar. We used a chromogenic agar (CHROMagar™ Staph aureus) specifically developed to identify *Staphylococcus aureus* as purple/pink colonies ^21^. *S. aureus* is of particular interest as it is the most frequent cause of infection in acute wounds ^27^. However, the Bactogram method could easily be applied to other types of chromogenic agar to visualize other bacteria of interest.

Figure 6 presents images of all CHROMagar plates from control wounds from the last 16 participants in the study where *S. aureus* was detected via MALDI-TOF in either the wound swab or the dressing fluid according to the data presented in Lundgren et al ^19^. The results suggest an accurate correspondence to MALDI-TOF results when using Bactogram with chromogenic agar, especially when *S. aureus* was identified in the wound swab (Subject A and B). For subject B on day 3, where no *S. aureus* was identified in the swab but was present in the dressing fluid, no evidence of this bacterium was seen on the Bactogram (pink/mauve colonies absent). For subjects C and D, *S. aureus* was only found in the dressing on day 8, and no clear sign of *S. aureus* was found on the CHROMagar plate on day 3. The red arrow for subject D, day 8, highlights an area with pink colonies that are not easily delimited (Figure 6) compatible with the detected presence of *S. aureus* ^19^.

**Figure 6:**
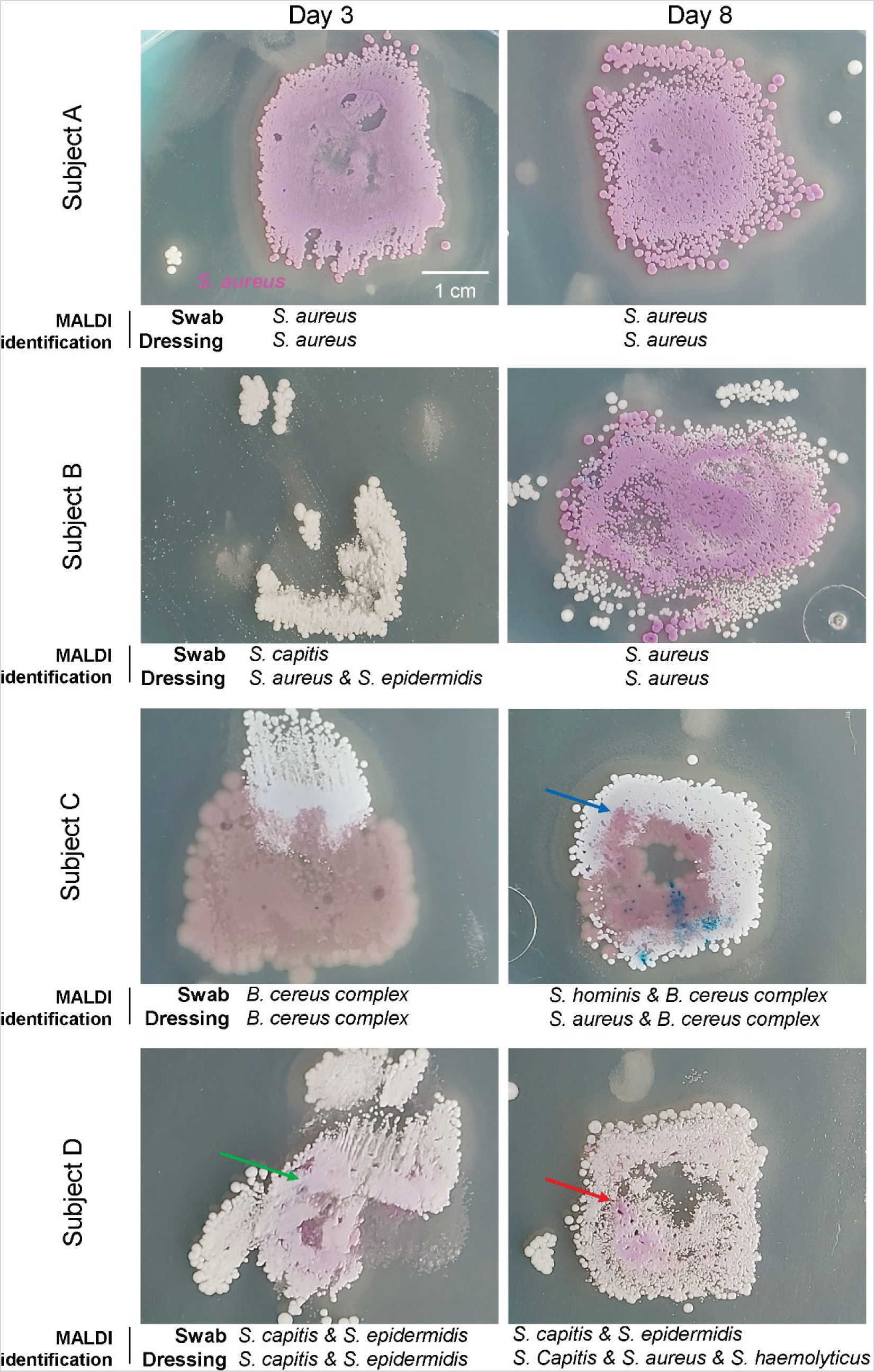
Proof-of-concept for Bactograms using chromogenic agar to visualize the growth of *Staphylococcus aureus*. As a proof of concept, a second Bactogram imprint was collected on a CHROMagar™ Staph aureus plate using the same filter paper after obtaining the imprint on the TH-agar plate on days 3 and 8 for the last 16 participants included in the study. The CHROMagar *S. aureus* plates specifically identify *S. aureus* as distinct colonies with a pink/mauve color. This figure presents photographs taken using an ordinary mobile camera of the resulting CHROMagar plates for all wounds where MALDI-TOF analysis showed growth of *S. aureus* in either the swab or dressing. The bacterial species identified by MALDI-TOF in that sample are specified under each image. The green arrow for subject D, day 3, highlights an area with an indistinct faint pink glow that does not indicate *S. aureus* according to the manufacturer’s instructions. The blue arrow for subject C, day 8, highlights an area with brown/pinkish undefined colonies that do not indicate *S. aureus* according to the manufacturer’s instructions. The red arrow for subject D, day 8, highlights an area with pinkish-looking colonies, suggestive of *S. aureus*.

However, the Bactogram method using chromogenic agar offers more than the presence or absence of *S. aureus*. The method also enables a direct visual evaluation of the spatial distribution and composition of cultivable bacterial species in the wound. As an example, Subject A’s wound bacteria appears to be dominated by a homogenous population of *S. aureus*, which populates the entire area under the dressing, with little variation observed between day 3 and day 8. In contrast, on day 3, subject B shows limited colonization, mainly in the lower right section of the dressing, consisting of bacterial species other than *S. aureus*. However, by day 8, there’s a noticeable surge in bacterial growth with both *S. aureus* and other bacteria together densely covering the whole wound and dressing area and even extending to the skin surrounding the dressing (Figure 6). Taken together, these data provide a proof-of-concept showing that the Bactogram method can enable spatial identification of *S. aureus* in a defined wound setting.

## 4. Discussion

This article presents a proof-of-concept of the Bactogram method, a novel method for spatial mapping of cultivable bacteria in wounds. We employed this method to study bacterial development in epidermal wounds made by the suction blister technique ^17^. Additionally, we developed two methods to quantify Bactograms based on manual visual scoring and computer-assisted image analysis. Together, these methods enabled us to visualize and quantify the spatial dynamics of bacterial colonization following a defined skin barrier damage. Large inter-wound differences were seen in the extent and location of bacterial colonization in the wound and under the dressing at all time points except directly after cleaning and wounding (day 1) where no bacterial growth was seen on either Bactograms or swabs. In spite of this, we still identified a common pattern of colonization where bacterial growth usually began on the skin underneath the dressing with bacteria most commonly migrating into the wound area in conjunction with the reepithelization of the wound ^26^. Computer-assisted Bactogram quantification in the wound and dressing area correlated moderately well with bacterial density (CFU/cm^2^) measured by CFU counts, the most common technique by which bacteria are studied in this setting.

There is an unmet need for new methods to study and assess bacteria in conditions involving skin barrier damage and subsequent colonization by potentially harmful bacteria, such as wound infections, atopic dermatitis, and chronic wounds ^6, 7, 9^. Available microbial sampling methods, such as swabs and tissue biopsies, provide a simplified picture of the bacteria since they usually only sample a small portion of the affected area without consideration of spatial differences. This oversimplification impedes the development of better strategies to prevent and treat these conditions as a high burden of bacteria may present without easily distinguishable clinical signs ^28^. Methods that enable the identification of areas with a high bacterial burden and/or pathogenic bacteria of specific interest are therefore important to be able to direct treatment to areas where the detrimental bacteria actually reside. The Bactogram method was developed in response to this unmet need and to address the size and spatial limitations of current microbiological sampling methods.

By studying Bactograms collected over time from each wound, we observed that bacteria developed in all wounds during the healing process. In general, bacteria first colonized the periphery of the wound, beneath the dressing and around the wound edges, and finally occupied the wound site on day 8, coinciding with the reepithelization of these wounds ^26^. However, this study also highlighted the variation between wounds, suggesting individual differences in the spatial extent of bacterial colonization. This finding aligns with existing research indicating that individuals possess distinct skin environments and microbiota ^10^. To our knowledge, this is the first comprehensive exploration of the spatial development of bacterial colonization in the same wounds over time from wounding to healing. Contrary to a widely accepted belief that bacteria primarily colonize the wound itself ^28–30^, our findings align with recent evidence published by Bay et al.^13^, and Le et al.^14^, which reports a predominance of bacteria residing at wound edges in acute and chronic wounds, respectively. A plausible physiological explanation for this observation could be that bacteria in the wound are directly exposed to the immune cells and antibacterial proteins and peptides present in wounds ^31–33^ which limits their growth. Wound edges are recognized as a crucial part of the clinical assessment of wound status ^15, 16^, suggesting that bacterial colonization in this area might affect wound outcomes. Therefore, if the wound edges are established as a possible bacterial reservoir it may be warranted to add more focus on bacterial management at wound edges in wound care routines. Additionally, the early and preferred colonization of the dressing area observed in this study could be a consequence of the dressing creating a suitable environment for bacterial growth. This is in line with other studies that note this phenomenon and the clinical importance of bacterial growth in and under dressings ^34, 35^. By being able to detect the preferred areas for bacterial growth, the Bactogram method offers a convenient methodology for future studies exploring bacterial colonization beneath the dressing and at wound edges and as a potential outcome measure to evaluate the effect of different dressings and wound treatments.

The comparison between the results from computer-assisted image analysis and conventional CFU counts in swabs and dressing samples showed that a relationship exists between bacterial coverage measured by the Bactogram method and the abundance of bacteria measured by a validated bacterial quantification method. However, the correlation was only moderately strong which makes sense in light of the different approaches used for quantification. CFU-counting of a swab sample quantifies the total amount of bacteria collected by the swab in an area by employing a method of serial dilutions, to get easily discernible and countable single colonies. In contrast, the Bactogram method lacks the possibility of serial dilution making it impossible to count colonies in most cases. The quantification has therefore to rely on quantifying the percentage of bacterial colonization in an area, which is an inherently different measure. As an example, two 1 mm^2^ areas with widely different numbers of bacteria collected by the filter paper, would reasonably both be equally likely to produce bacterial impression on the agar plate, even though their bacterial densities differ significantly, and would therefore appear similarly on the Bactogram and result in the same percentage of bacterial colonization. In line with this, our results demonstrate better concordance of the methods at lower bacterial levels, where the saturation effect of the Bactogram method is less pronounced. Accordingly, when the swab reveals no CFUs the Bactogram usually also exhibits no colonies. A few exceptions were noted where colonies were detected on the Bactogram but not by the swab, and the other way around. However, a potential explanation for this is the challenge of determining whether sparse bacterial colonies on the Bactogram are associated with the wound or the dressing. This ambiguity may be more prevalent in wounds with few bacteria, such as some of the wounds in this study, and less evident in clinical scenarios with higher bacterial colonization, like chronic wounds ^9^, burn wounds ^36^, or atopic dermatitis ^37^. In this study, it appears that at least approximately 3 × 10^3^ CFU/cm^2^ bacteria collected by the swab and approximately 5 × 10^3^ CFU/cm^2^ collected in the dressing fluid are required before colonies become visible on the Bactogram. This suggests that the currently used Bactogram method cannot consistently identify bacteria at levels below approximately 5 × 10^3^ CFU/cm^2^. This limitation may arise from multiple factors, including whether the bacteria are in a planktonic or biofilm growth state and whether they are primarily located superficially or deep within the skin and wound. In comparison, the swabbing method uses a stronger and more localized pressure probably enabling the collection of slightly deeper-residing bacteria. However, if the Bactogram method would capture substantially lower levels of bacteria, it might also lead to the collection of less clinically relevant colonization due to the omnipresence of bacteria on the skin ^2^. Moreover, the more uniform pressure used in the Bactogram method might also enable a more reproducible bacterial collection as the Bactogram is less prone to differences caused by small variations in the exact area sampled and the amount of pressure applied when swabbing. Interestingly, the detection limit of the commercially available wound imaging device MolecuLight, claims to detect clinically significant amounts of bacteria with a detection threshold of bacterial levels above 10^4^ CFU/gram of tissue ^14^.

Taken together, the Bactogram method presents a promising addition to traditional bacterial sampling techniques like swabs or biopsies. It not only covers a larger sampling area (120 vs 1 cm^2^) but, unlike conventional methods, also allows for detailed spatial analysis of bacterial distribution. Moreover, our data demonstrates that the Bactogram method can produce a semi-quantitative analysis of the presence and distribution of cultivable bacteria at various anatomical locations, without needing the labor-intensive plating and manual counting associated with traditional methods.

However, it is essential to consider several constraints associated with methodology and our results. Growth rates vary between bacterial species ^38^ and our measurement of bacterial coverage after a fixed time in one growth condition may therefore preferentially detect bacteria that are faster-growing or that produce large colonies. However, this challenge is not unique to the Bactogram method and is also present in conventional methods such as swabs and quantitative bacterial counts ^39^. Another possible limitation of the Bactogram method concerns the propensity of bacteria to adhere to the filter paper during collection and their likelihood to transfer from the filter paper to the agar plate. Differences in bacterial adherence and transfer may be a function of bacterial species and their planktonic or biofilm growth states, which could be important for some applications of this method and thus warrants further investigation in upcoming studies. Nonetheless, this limitation is not unique to the Bactogram method and can also apply to swabs, where the method’s success also depends on the bacteria’s attachment properties ^40^. Although the Bactogram method presents certain limitations, it is a new tool for investigating the spatial distribution of bacteria. The utility of Bactogram is also highly dependent on the growth medium used. Different bacterial species require specific growth media ^39^, raising the possibility of overlooking certain bacteria with unique growth requirements. Future applications of the method must carefully consider the choice of growth medium. A potential improvement might involve capturing multiple Bactograms from the same wound sample and cultivating them on different plates to obtain a more comprehensive bacterial profile. We have shown a proof of concept for collecting a second bacterial imprint on a chromogenic agar to identify *S. aureus*, highlighting this as an area for future development of the method. Though the data were limited, it showed a promising correspondence with the results of bacterial identification using MALDI-TOF analysis. *S. aureus* is the most common pathogen overall in skin and wound infections ^27, 41^. Furthermore, S. *aureus* colonization is a known risk factor and cause for surgical site infections (SSIs) after full-thickness skin grafting ^42^ and impaired wound healing ^43^. Further studies are warranted as easy identification of areas with bacteria such as *S. aureus* could provide clinicians with a spatial understanding of colonization patterns and aid in early bacterial detection, enabling early interventions aiming to reduce wound infections.

## 5. Conclusion

In conclusion, the Bactogram method presents a new approach that addresses some of the limitations of current bacterial sampling methods, by offering a straightforward technique for visualizing the spatial distribution of cultivable bacteria across a large area. By applying this method on suction blister wounds, we described the spatiotemporal development of bacteria following epidermal wounding, showing that bacteria resides predominantly under the dressing and near wound edges during re-epithelization, with substantial inter-individual variations in the spatial extent of bacterial colonization. Collectively, these observations call for a better understanding of the spatial distribution of bacteria in diseases with skin barrier damage, which we believe to be fundamental in propelling future research and development of effective treatment strategies.

## Author Contributions

The following authors contributed to each of the following roles (defined according to the CRediT taxonomy): Conceptualization, KW, MP, AS; design of methodology, KW, FF, GP, MP, AS; formal analysis (statistics), KW, FF, JF; investigation (performing experiments and data collection) and validation, KW, FF, SL, JC, GP, ACS; resources (provision of subjects and samples), KW, SL, KS; data curation, KW, FF, JF; Software, KW, FF; writing (original draft preparation), KW, FF, JF; writing (review and editing), KW FF, SL, JF, JC, GP, ACS, KS, MP, AS; visualization KW, FF; supervision, KW, AS; project administration, KW, JF, AS; funding acquisition, AS. All authors have read and agreed to the published version of the manuscript.

## Data Availability

The generated pictures of the Bactograms will be available on reasonable request, after the publication of the treated wound data in a separate publication, since the original images inevitably include both treated and untreated wounds. The code used for the ImageJ macros can be found in Supplementary Data 3. The Python code for creating spatial heat maps can be found in Supplementary Data 4. The complete score data from both the visual scoring and the computer-assisted method is provided in Supplementary Data 5. Further information or instructions required to reanalyze the data reported in this paper are available from the lead contact upon reasonable request.

## Acknowledgments

The exploratory data presented here was supported by grants from the Swedish Research Council (project 2017-02341, 2020-02016), Edvard Welanders Stiftelse and Finsenstiftelsen (Hudfonden), the Royal Physiographic Society, the Crafoord and Österlund Foundations, and the Swedish Government Funds for Clinical Research (ALF). Xinnate AB provided the project management resources and expertise for the regulatory development enabling the clinical parts of the Safety study that generated the control samples used in this work. We thank Susanne Erdmann and Anne Nielsen and other personnel at the Department of Dermatology Lund and the Clinical Trial Unit at Skane University Hospital Lund for support.

## Declaration of interests

A.S. is a founder of in2cure AB, a parent company of Xinnate AB, companies that are developing therapies based on thrombin-derived peptides and variants. G.P. is employed part-time (20%) by Xinnate AB. The other authors have declared that no conflict of interest exists.

## Supplementary materials

Figure 1 – Examples of smeared Bactograms from the eight first subjects.

Figure 2 – Screenshot from ImageJ when using the macro to calculate bacterial coverage in the dressing area.

Figure 3 – The decision tree used to determine how to align the square in ImageJ.

Figure 4 – Sensitivity analysis with smeared images included.

Figure 5 – Comparison between quantification methods.

Figure 6 – Visual description of the method used to create the spatial heat maps.

Data 1 – The guide used by the raters doing the visual scoring.

Data 2 – Image used to determine the size of the dressing area.

Data 3 – Instructions and code used for the computer-assisted quantification.

Data 4 – Python code for the spatial heatmap.

Data 5 – Data used in the manuscript.

## Notes

### Clinical Trial

NCT05378997

### Clinical Protocols

https://doi.org/10.1136/bmjopen-2022-064866

### Author Declarations

Etikprövningsmyndigheten (Swedish Ethics committee) gave ethical approval for this work.

