## Supplementary Data 1 for "Bactogram: Spatial Analysis of Bacterial Colonization in Epidermal Wounds"

### Wound

Try to find a wound area the size of the red circle in the examples and assess the amount of bacteria in that area.

**Wound 0**

When the wound is empty, or very close to empty

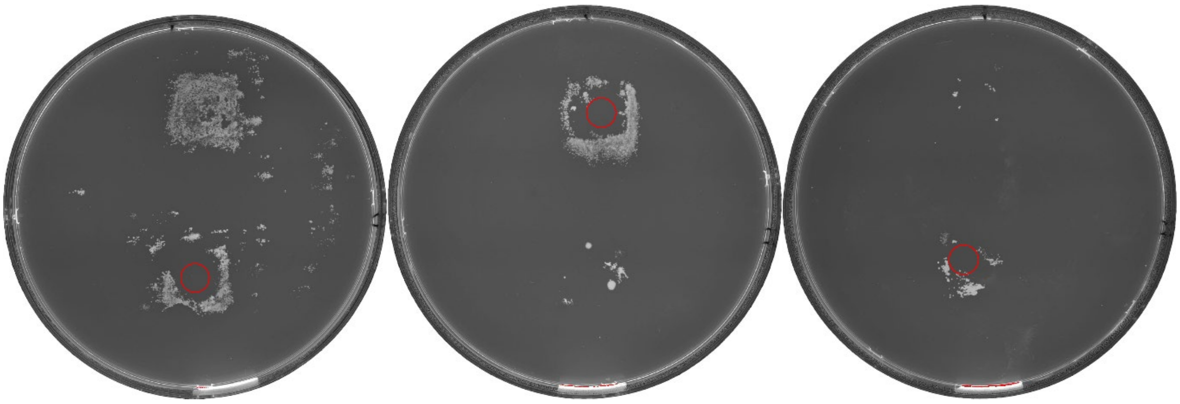

**Wound 1**

When the wound is almost empty, but there are visual traces of bacteria

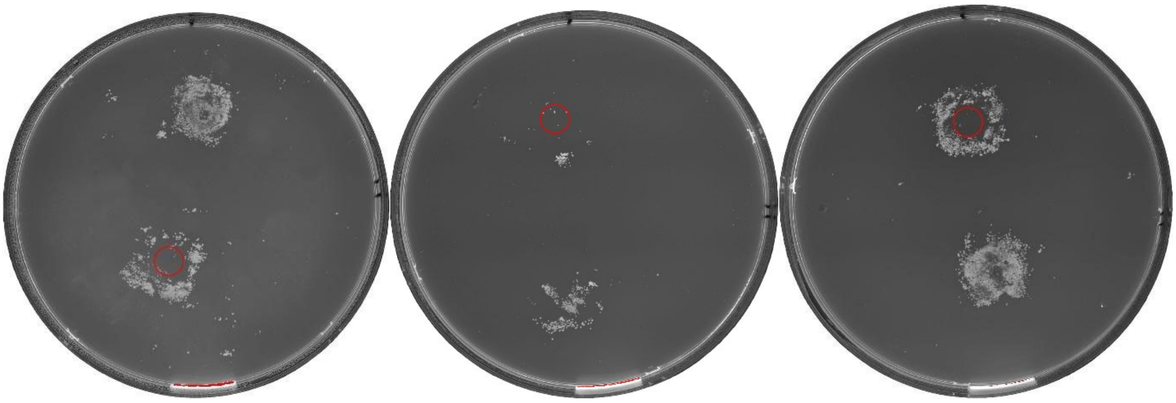

**Wound 2**

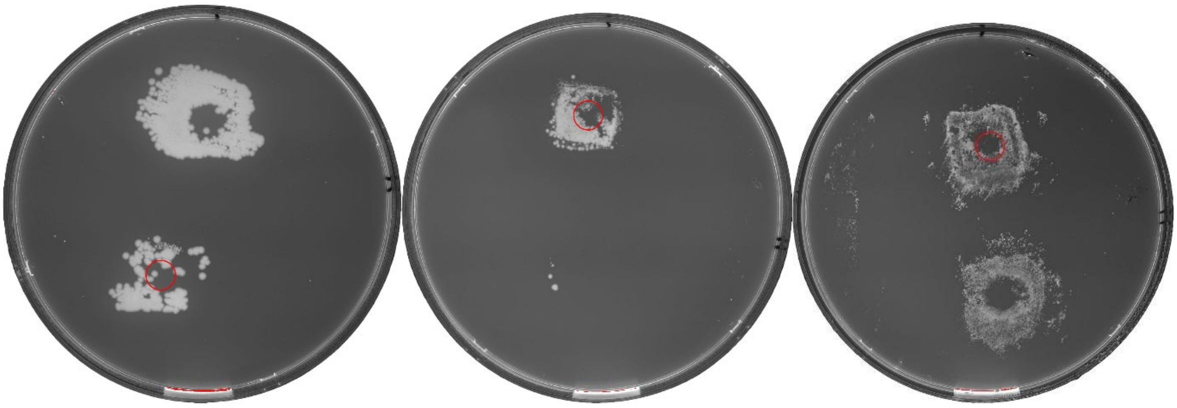

**Wound 3**

When the area is close to full, but not completely

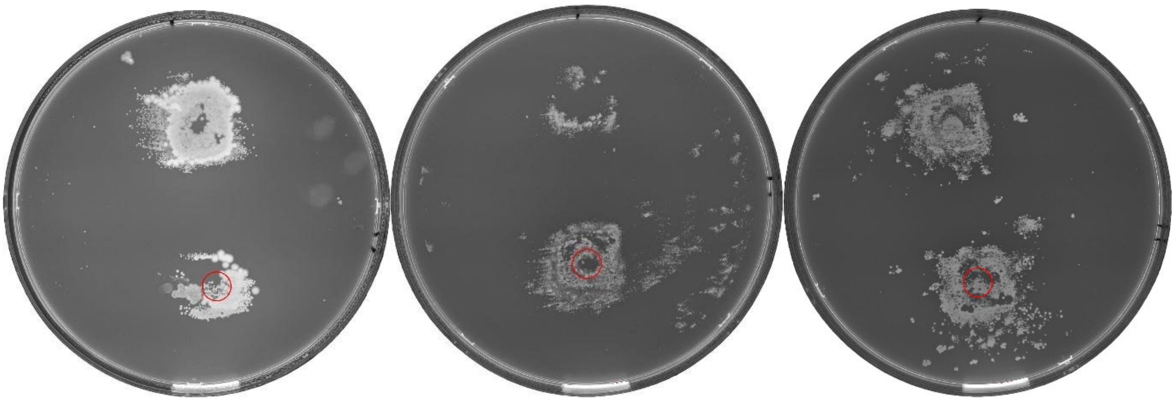

**Wound 4**

When the wound area is completely full

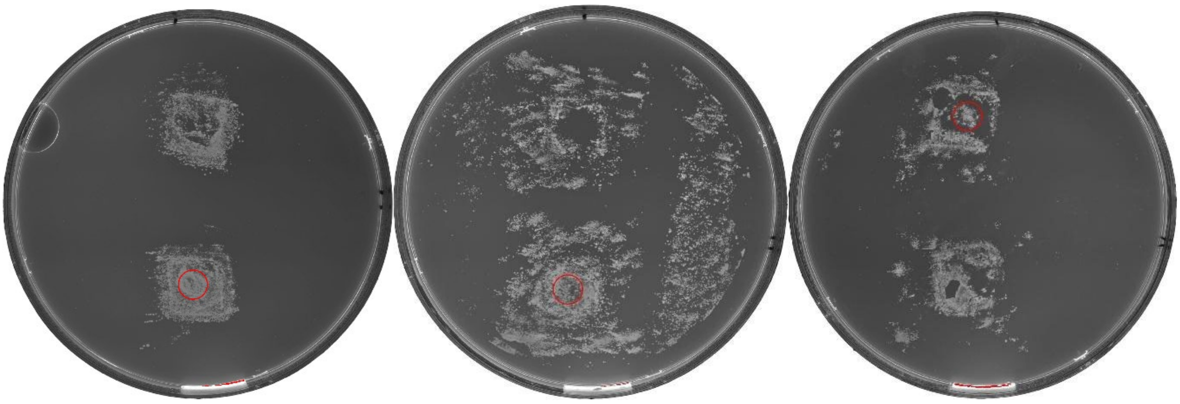

### Dressing

Try to find an area the size of the red square that corresponds to the dressing area and assess the amount of bacteria that are in that area, but that are not in the wound described above. It can sometimes be hard to find a good area to look for, then it can be helpful to compare to the other wound since the distance between the wounds should always be the same. *Remember to neglect the bacteria in the wound.*

**Dressing 0**

Empty or very close to empty

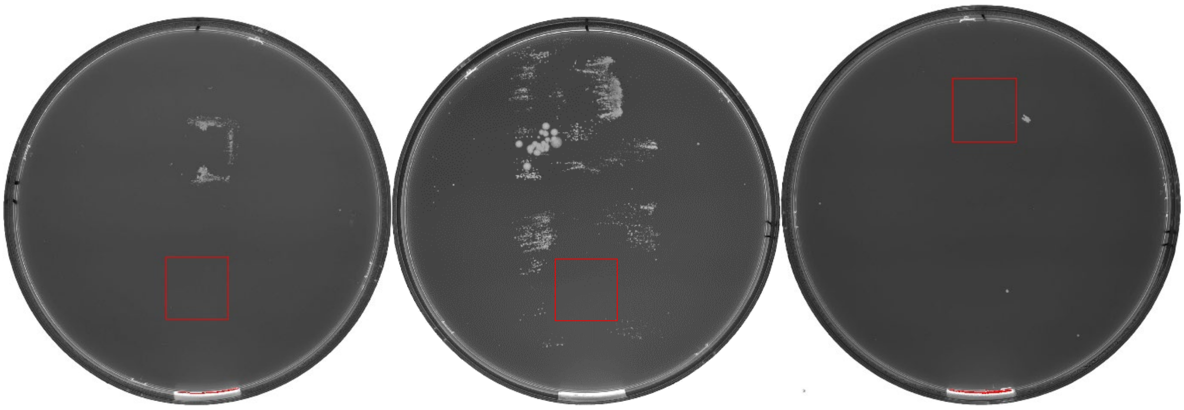

**Dressing 1**

Low amounts of bacteria, but not empty

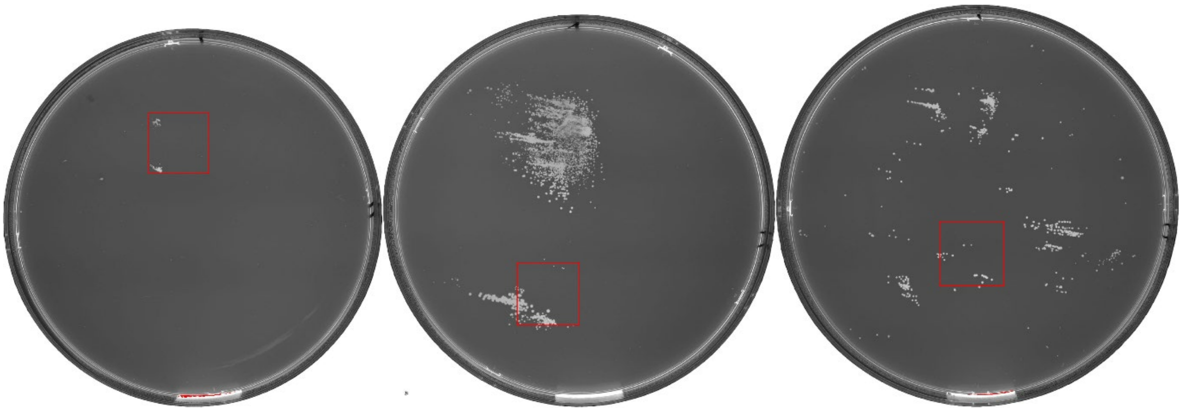

**Dressing 2**

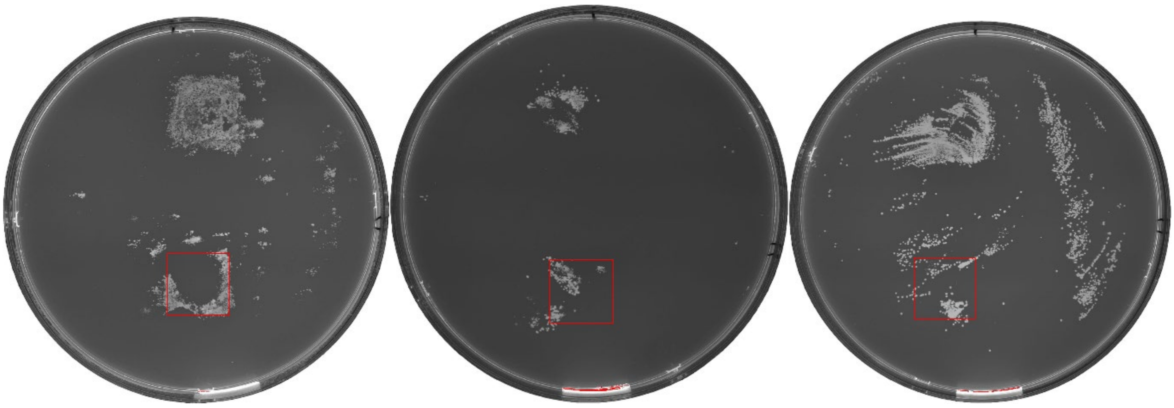

**Dressing 3**

When the bacteria cover most of the outer part of the dressing area, but not fully

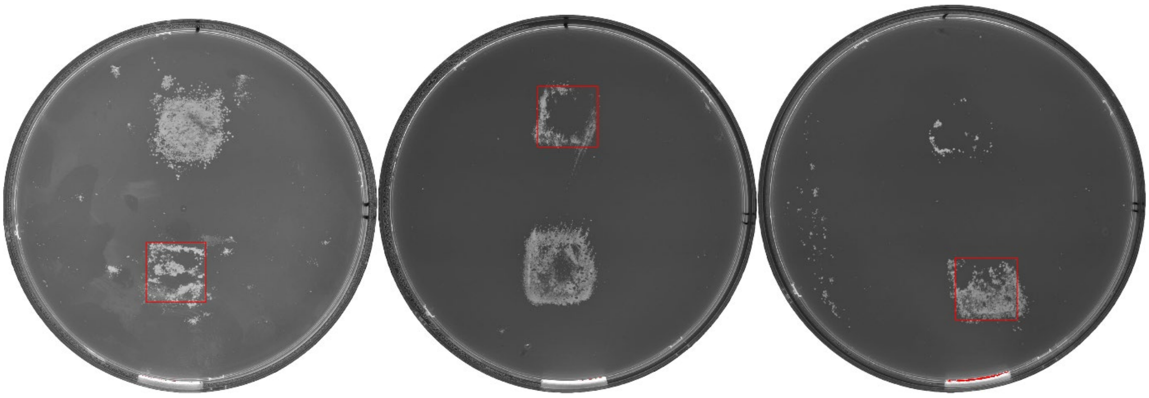

**Dressing 4**

When the bacteria cover all sides and the inner part (center not included as can be seen in the first and last example of Dressing 4)

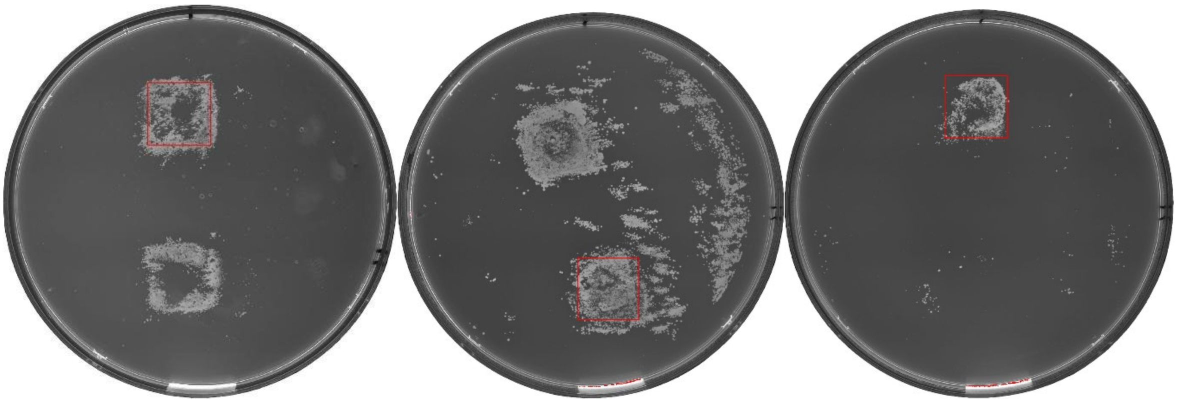

### Skin

Look at the bacteria that are not covered by the red square in the dressing section. When bacteria are very remote from the dressing area, it is up to you to decide if they belong to the top or bottom wound by looking at proximity.

**Skin 0**

Almost everything that belongs to the wound is in the square

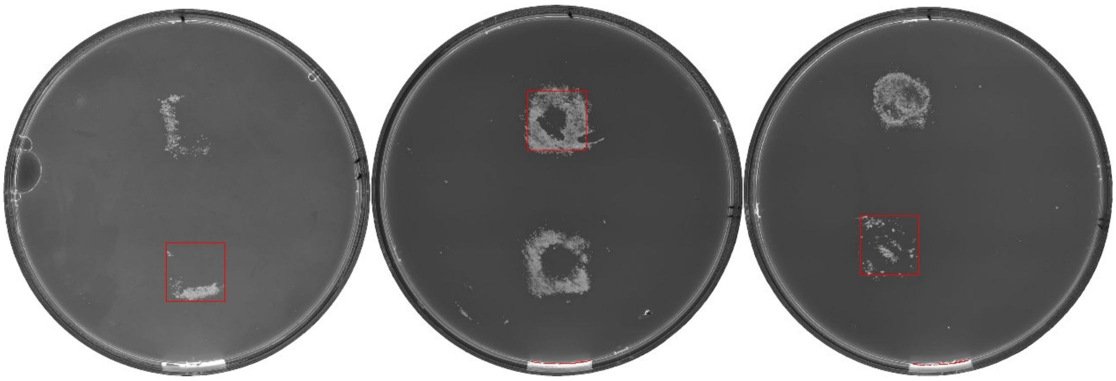

**Skin 1**

When there are some bacteria that are not in the square, but especially when the amount of bacteria outside of the square is close to as much as in the square (example 3 of skin 1)

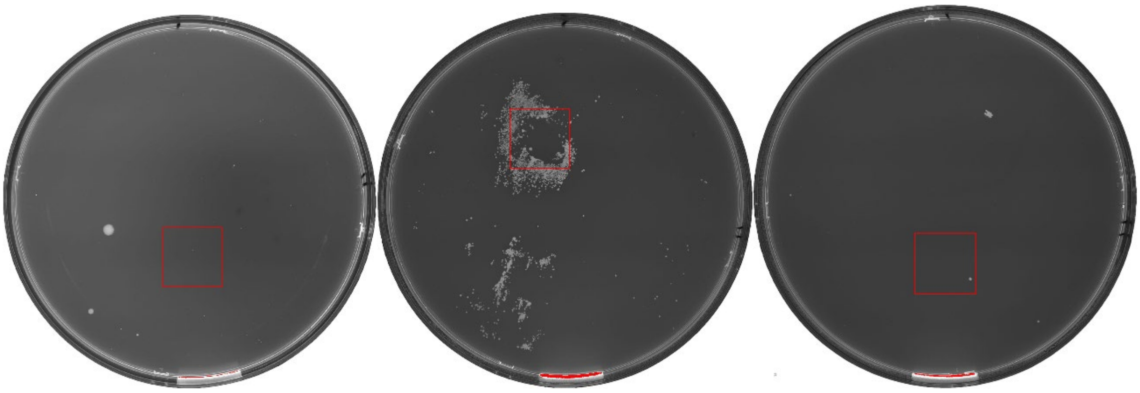

**Skin 2**

Similar amount of bacteria as in skin 1, but more spread out from the dressing area

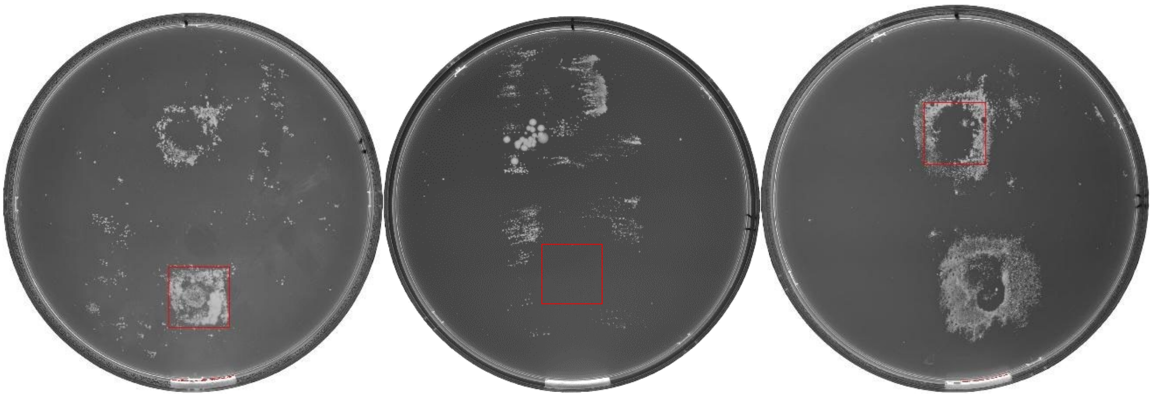

**Skin 3**

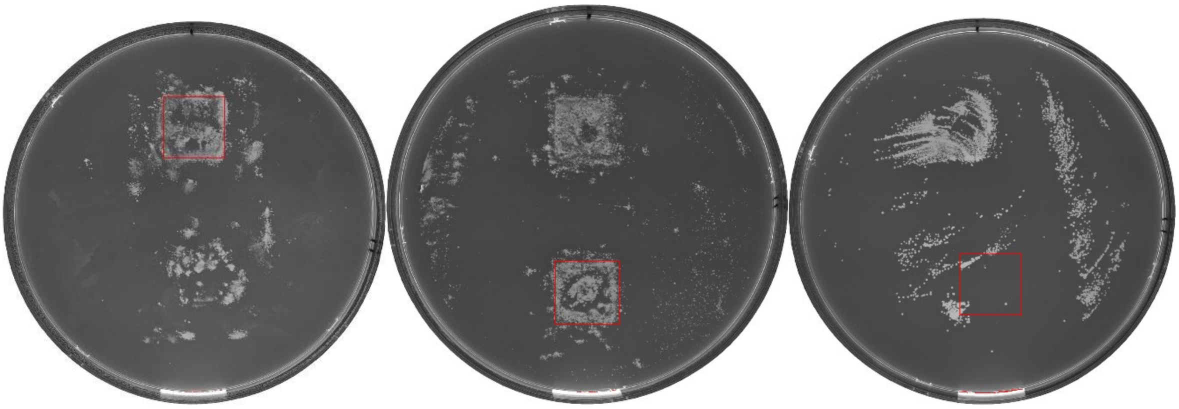

**Skin 4**

Bacteria outside of square cover most of the plate

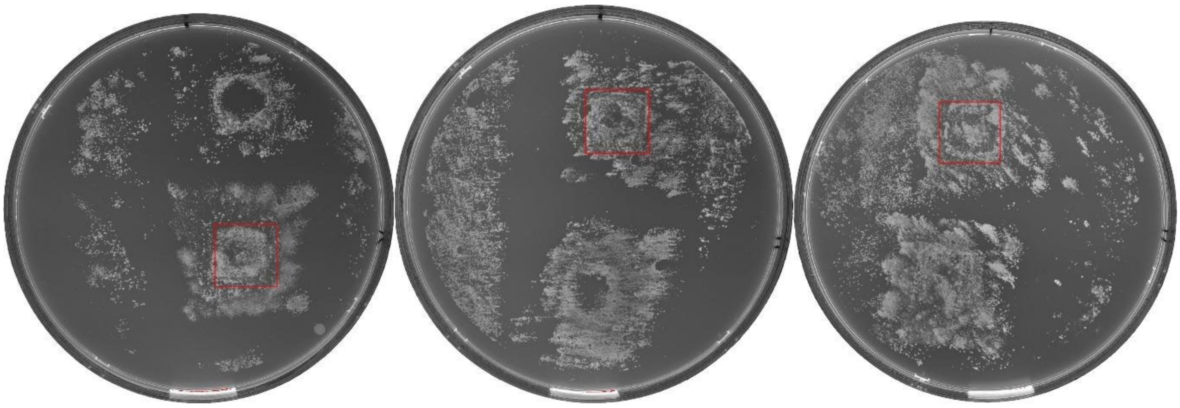
