## Supplementary figures and images for "Bactogram: Spatial Analysis of Bacterial Colonization in Epidermal Wounds"

### Supplementary Data 2

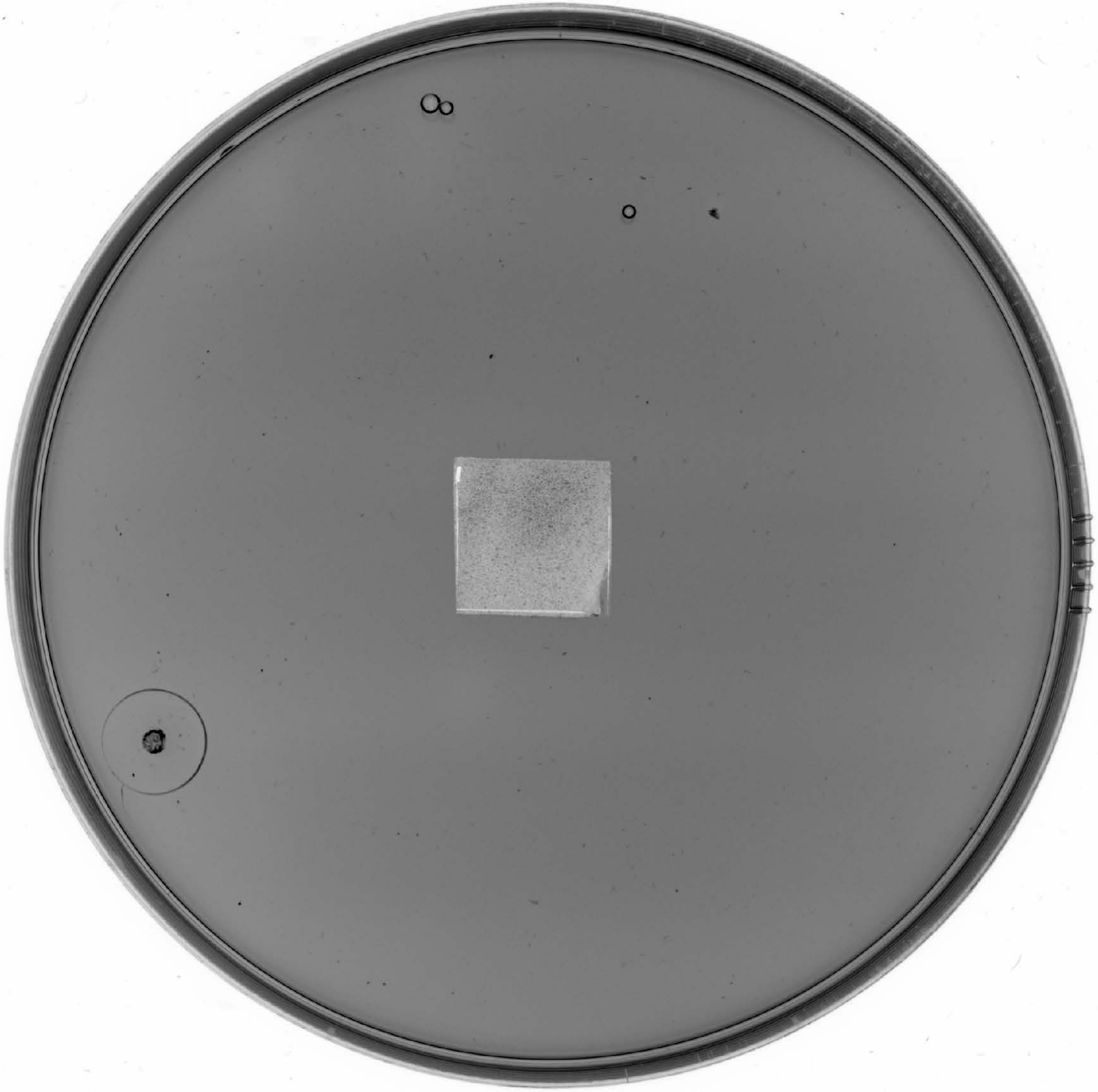

### Supplementary Figure 1

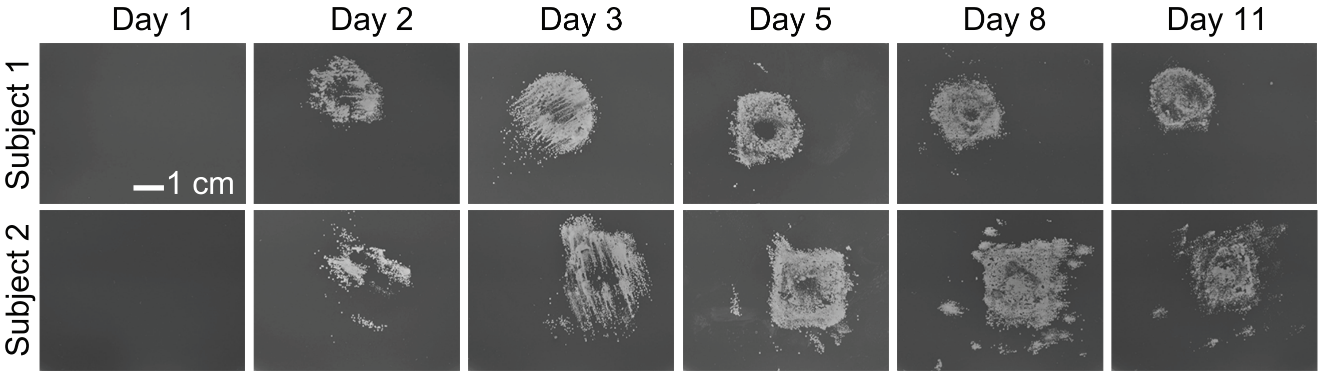

### Supplementary Figure 2

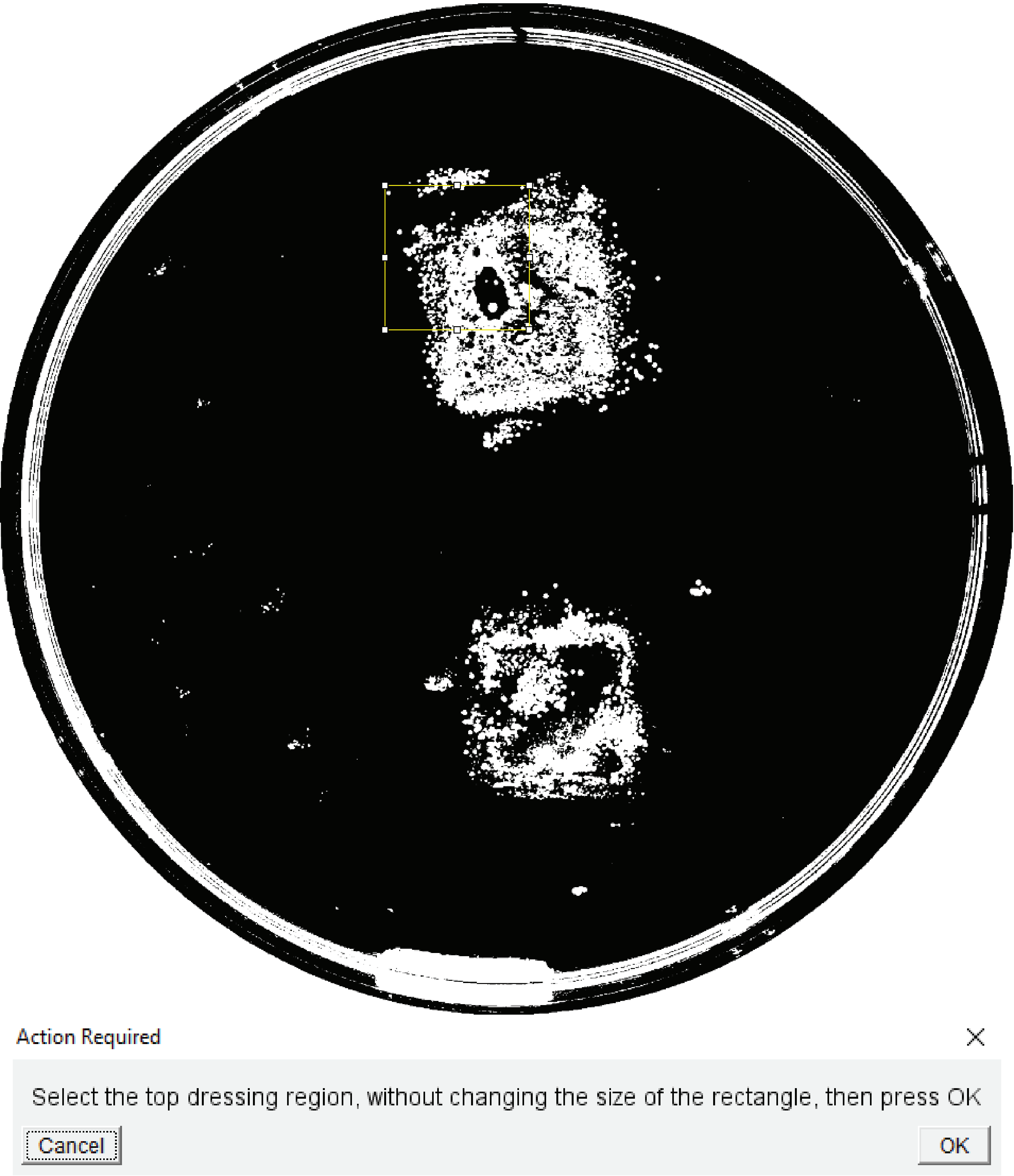

### Supplementary Figure 3

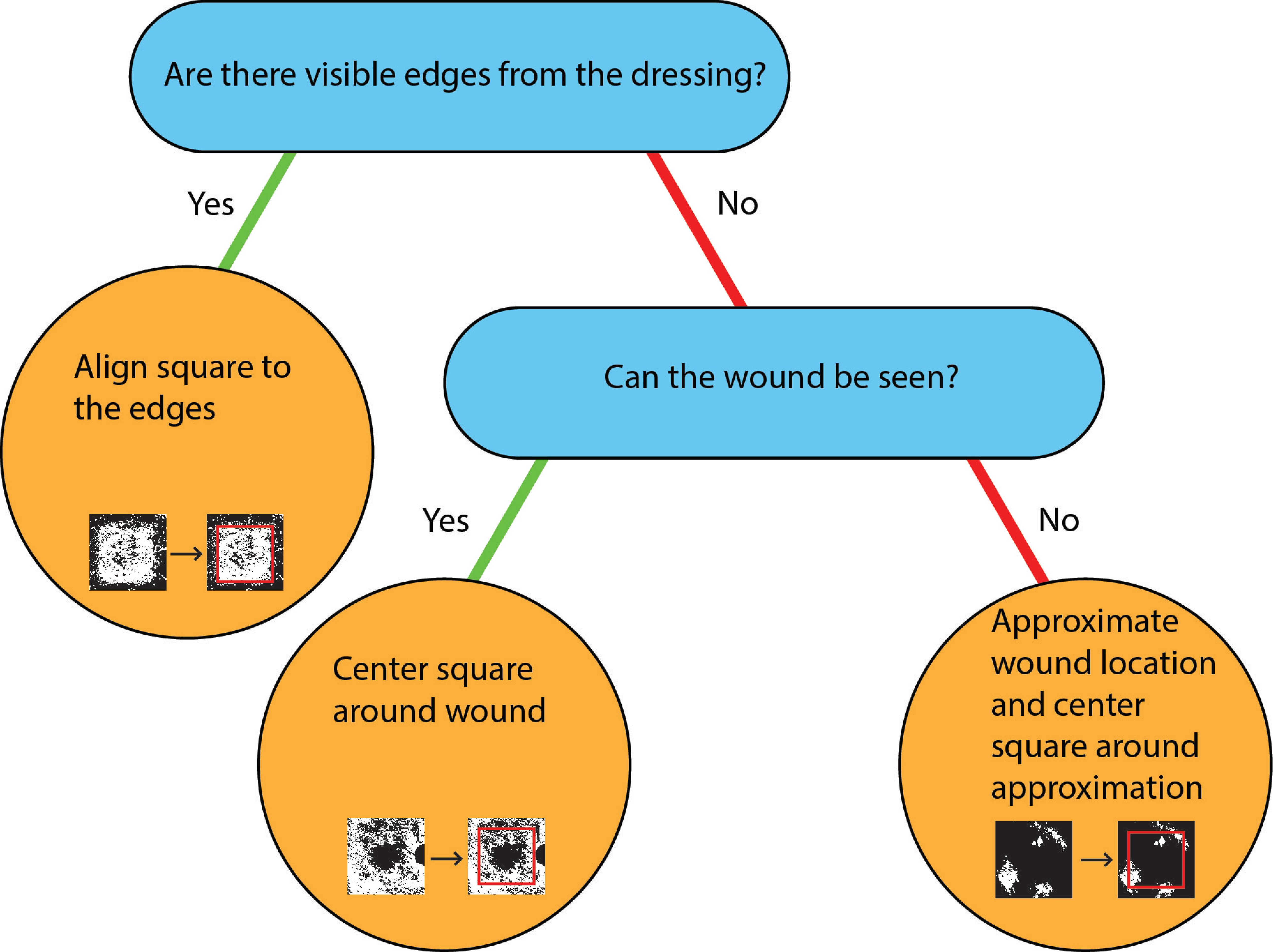

### Supplementary Figure 4

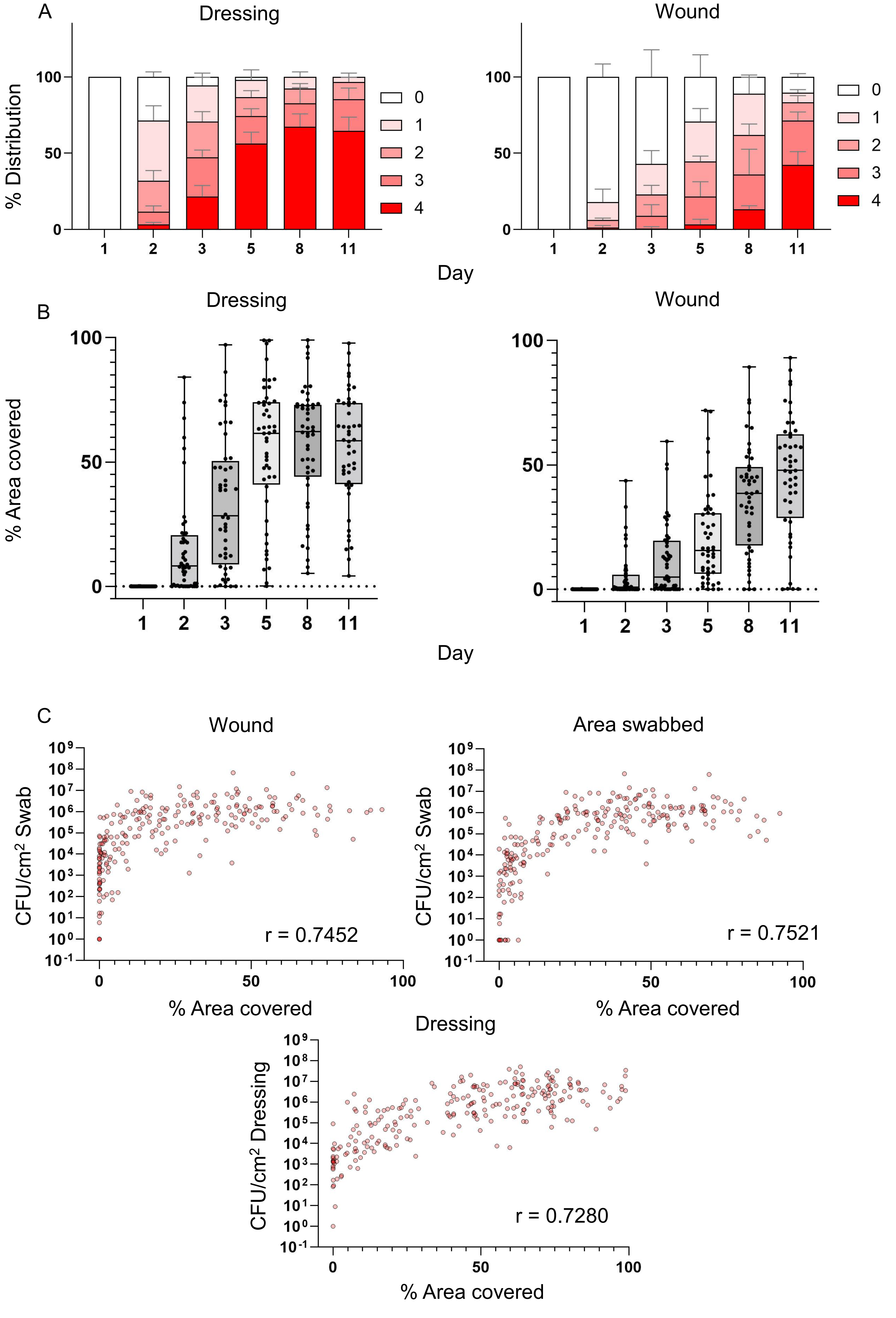

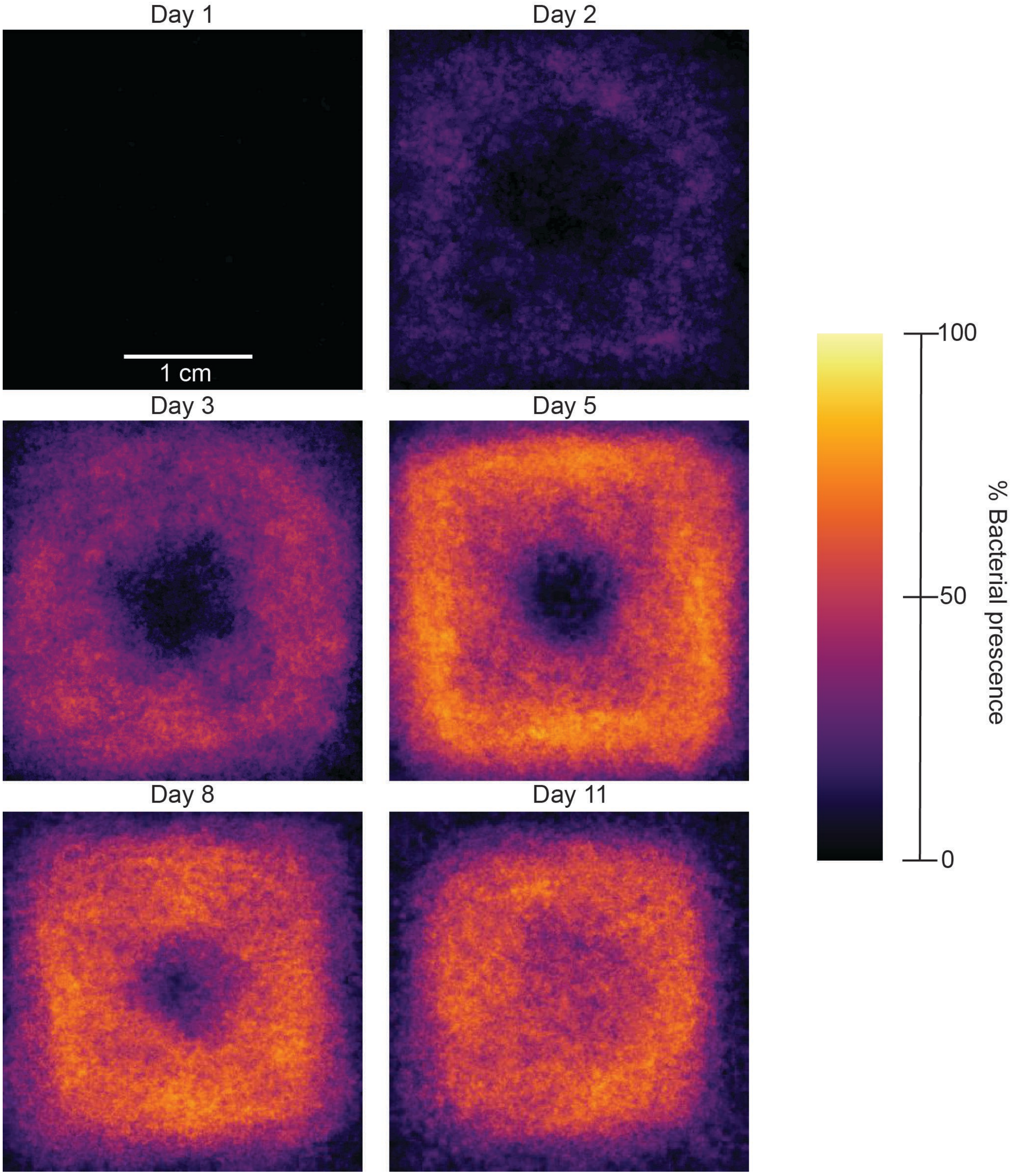
