## Supplementary Data 3 for "Bactogram: Spatial Analysis of Bacterial Colonization in Epidermal Wounds"

### ImageJ measurement instructions.

- Open ImageJ and go to ”Plugins” → ”Macros” → ”Startup Macros”.
- Add the following macros to the end of the page in the “Startup Macros” pop-up:

```
macro "MeasureMacroRectangle [f1]" {
	setTool("Rectangle");
	makeRectangle(669, 321, 250, 250);
	waitForUser("Select the top dressing region, without changing the size of the rectangle, then press OK");
	run("Measure");
	makeRectangle(669, 1041, 250, 250);
	waitForUser("Select the bottom periwound region, without changing the size of the rectangle, then press OK");
	run("Measure");
}
macro "MeasureMacroCircle [f2]" {
	setTool("Oval");
	makeOval(756, 435, 125, 125);
	waitForUser("Select the top wound region, without changing the size of the circle, then press OK");
	run("Measure");
	makeOval(750, 1176, 125, 125);
	waitForUser("Select the bottom wound region, without changing the size of the circle, then press OK");
	run("Measure");
}

macro "MeasureMacroCircleExtended [f3]" {
	setTool("Oval");
	makeOval(756, 435, 175, 175);
	waitForUser("Select the top wound region, without changing the size of the circle, then press OK");
	run("Measure");
```

```
        makeOval(750, 1176, 175, 175);

        waitForUser("Select the bottom wound region, without changing the size of the circle, then press OK");

        run("Measure");

}
```

- Restart ImageJ.
- Go to “Analyze” → “Set Measurements” and make sure that the boxes named “Area fraction” and “Integrated density” are ticked.
- Go to “File” → “Import” → “Image sequence”. Choose the folder containing the binary Bactogram images that are to be analyzed.
- To analyze the square area, click f1. Follow the instructions in the prompt. Go to the next image in the image sequence and repeat.
- To analyze the wound area, click f2. Follow the instructions in the prompt. Go to the next image in the image sequence and repeat.
- To analyze the swabbed area, click f3. Follow the instructions in the prompt. Go to the next image in the image sequence and repeat.
