## Supplementary Data 4 for "Bactogram: Spatial Analysis of Bacterial Colonization in Epidermal Wounds": Supplementary Data 4 - Create heatmap images.py

```
import numpy as np
import imageio as ii
import os
from PIL import Image as im
import matplotlib.pyplot as plt


# Take a string and add all 350x350 TIF-images that includes that string in its filename to one
# Calculate the average intensity for each pixel
# Return this as an image
def avgDayImg(day):
    totImg = np.zeros((350,350), dtype=int)
    imgCount = 0
    for x in os.listdir():
        if x.endswith(".tif"):
            if day in x:
                img = ii.imread(x)
                totImg = totImg+img
                imgCount = imgCount+1
    avgImg = totImg/imgCount
    return im.fromarray(avgImg)
    

# Apply a colormap to a grayscale image
# Return this as a matrix
def turnGrayScaleToColor(colorMap, imgMatrix):
    endMatrix = np.zeros((350,350,3))
    for x in range(0, 350):
        for y in range(0, 350):
            for z in range(0, 3):    
                endMatrix[x,y,z] = colorMap(round(imgMatrix[x,y]))[z]*255
    return endMatrix


# Convert a color matrix into an image and save it with the given filename. 
def saveHeatMap(colorMatrix,fileName):
    colorImage = im.fromarray(colorMatrix.astype(np.uint8))
    colorImage.save(fileName)


# Set the current working directory to the directory containing the 350x350 pixel binary images
workingFolder = "XX"
os.chdir(workingFolder)


# Create grayscale images
d1ImgGS = avgDayImg("-d1-")
d2ImgGS = avgDayImg("-d2-")
d3ImgGS = avgDayImg("-d3-")
d5ImgGS = avgDayImg("-d5-")
d8ImgGS = avgDayImg("-d8-")
d11ImgGS = avgDayImg("-d11-")


# Choose colormap to be used for heatmaps
colorMap = plt.get_cmap("inferno")


# Create color matrices and save as jpg-files
d1ColorMatrix = turnGrayScaleToColor(colorMap, np.asarray(d1ImgGS))
saveHeatMap(d1ColorMatrix,"Day 1.jpg")

d2ColorMatrix = turnGrayScaleToColor(colorMap, np.asarray(d2ImgGS))
saveHeatMap(d2ColorMatrix,"Day 2.jpg")

d3ColorMatrix = turnGrayScaleToColor(colorMap, np.asarray(d3ImgGS))
saveHeatMap(d3ColorMatrix,"Day 3.jpg")

d5ColorMatrix = turnGrayScaleToColor(colorMap, np.asarray(d5ImgGS))
saveHeatMap(d5ColorMatrix,"Day 5.jpg")

d8ColorMatrix = turnGrayScaleToColor(colorMap, np.asarray(d8ImgGS))
saveHeatMap(d8ColorMatrix,"Day 8.jpg")

d11ColorMatrix = turnGrayScaleToColor(colorMap, np.asarray(d11ImgGS))
saveHeatMap(d11ColorMatrix,"Day 11.jpg")
```
