## Supplementary Figure 5 for "Bactogram: Spatial Analysis of Bacterial Colonization in Epidermal Wounds"

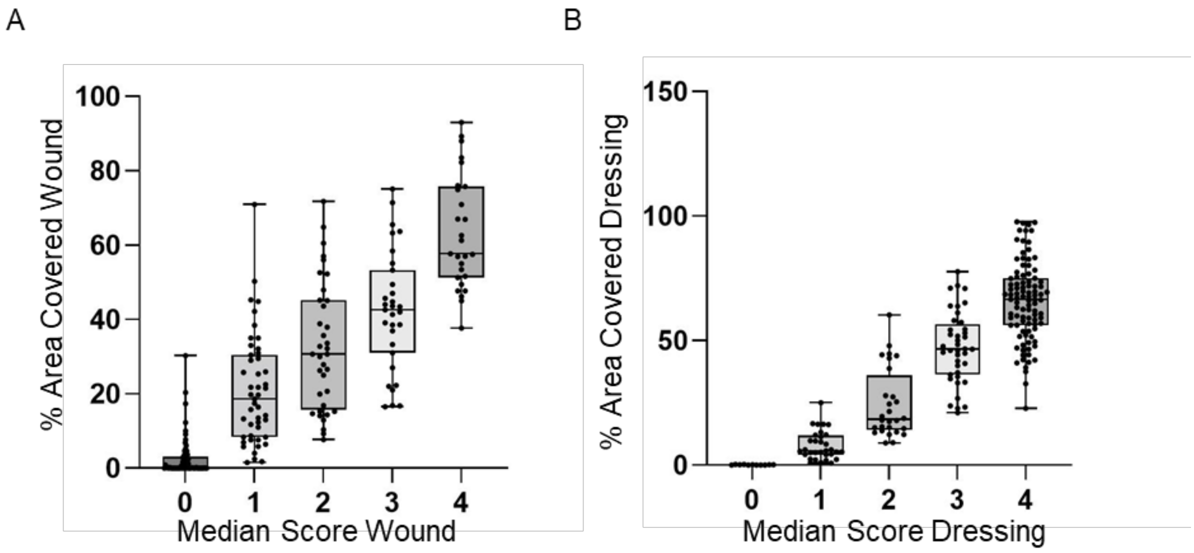


**Supplementary 5:** Comparison of the two quantification methods

To determine whether the two scoring methods yield similar information about bacterial coverage, we plotted the bacterial coverage quantified by ImageJ-assisted quantification in each median score category for both the wound (A) and dressing (B) area. The correlation was also quantified using Spearman's correlation coefficient. The two methods were positively correlated for both the wound ($r=0.8623$, $p<0.0001$) and dressing area ($r=0.8394$, $p<0.001$).
