## Supplementary Figure 6 for "Bactogram: Spatial Analysis of Bacterial Colonization in Epidermal Wounds"

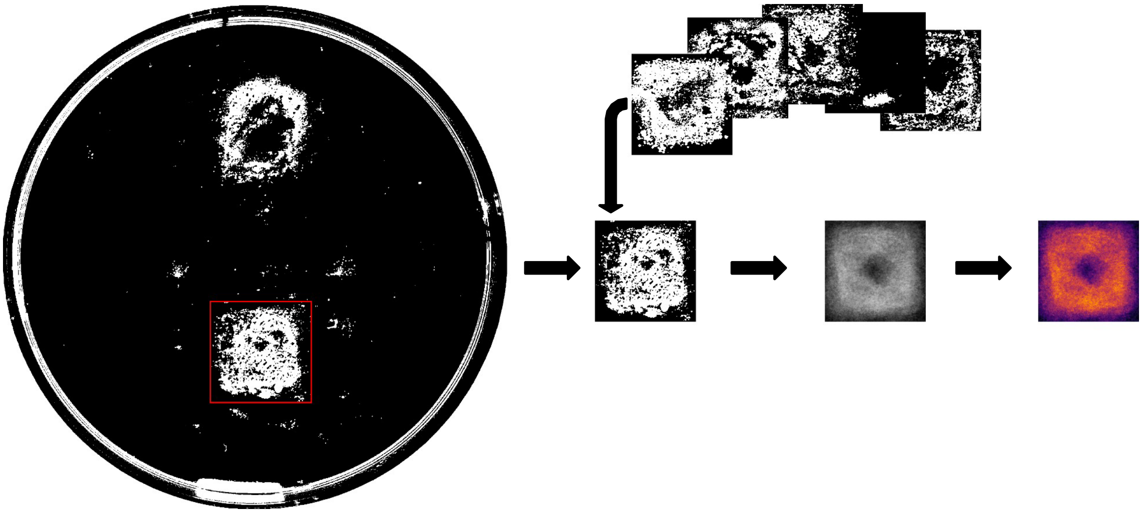

**Supplementary 6:** Visual description of the spatial heatmaps of bacterial presence.

To investigate the most common locations for bacterial growth in and around the wound, spatial heat maps were created for each day. To create the spatial heat map, the binary Bactogram images were used. Firstly, we cropped 350 x 350 pixel squares centered around the dressing area. These cropped images were grouped by time point and overlapped on each other. Using a Python script, we summed all overlapping pixels (0 no colony, 1 colony), yielding a greyscale visualization. To get the percentage of bacterial presence, we divided the pixel values in these grayscale images by the total number of overlapping pictures. This percentage was then used to color the heat map according to the color scale, where a lighter, more yellow color signifies a higher percentage of all Bactograms having bacterial coverage at that particular location.
